# Positive attribute framing increases COVID-19 booster vaccine intention for unfamiliar vaccines

**DOI:** 10.1101/2022.01.25.22269855

**Authors:** K. Barnes, B. Colagiuri

**Affiliations:** University of Sydney, Australia

## Abstract

Positive framing has been proposed as a potential intervention to increase COVID-19 vaccination intentions. However, most available research has examined fictitious or unfamiliar treatments. This pre-registered study compared positively and negatively attribute-framed side effect information for real COVID-19 booster vaccines (AstraZeneca, Pfizer, Moderna) and measured booster intentions pre- and post-intervention in 1,222 UK-based participants. As hypothesised, vaccine familiarity significantly modulated the effect of framing. While positive framing was effective for the least familiar vaccine (i.e., Moderna), standard negative framing appeared to increase intentions for familiar vaccines (AstraZeneca/Pfizer), particularly among those with low baseline intentions. These findings provide important new evidence that positive framing could improve vaccine uptake globally when switches or new developments require individuals to receive less familiar vaccines – as is currently the case for millions of booster vaccines across the world. Positive framing of familiar vaccines, however, should be treated with caution.

## Introduction

With vaccine efficacy for Severe Acute Respiratory Syndrome Coronavirus 2 (COVID-19) waning over time^1,2^ and reduced for emerging variants^3,4^, many countries are accelerating their COVID-19 booster programmes^5,6^. However, vaccine availability does not necessarily translate to vaccine acceptance^7^, with the World Health Organization (WHO) recognising vaccine hesitancy as a global health threat^8^. Side effect apprehension is a primary factor driving hesitancy^9^, with 90% of COVID-19 vaccine refusers fearing side effects more than COVID-19 itself^10^. Further, side effect severity from initial doses has been associated with booster vaccine hesitancy^11^. Reducing perceptions of side effects appears vital for increasing booster acceptance and reducing the global burden COVID-19.

The WHO^12^ has suggested the framing of vaccine-relevant information^13^ could provide a method of reducing negative perceptions. Positive attribute framing, where side effect information is framed in terms of the inverse incidence rate (e.g., “60% will ***not*** get a sore arm”) as opposed to typical negative framing with the standard incidence rate (e.g., “40% will get a sore arm”) could be particularly useful for combatting COVID-19 vaccine hesitancy. First, it is directly applicable to side effects. Second, informed consent is maintained due to statistical consistency across frames^14^. Third, there is preliminary evidence that positive attribute framing can improve vaccination attitudes in other settings. For example, one study on the influenza vaccine found positive attribute framing (hereafter positive framing) reduced the expectation and experience of side effects, increased perceived protection from influenza, and reduced distortions in the perception of side effect risk^15^, with results replicated for other vaccine types^16^.

The handful of studies examining framing on COVID-19 vaccine intention have produced mixed results^17-19^. However, those studies focused on vaccine-naïve individuals, did not employ attribute framing, and did not concern booster intentions. Further, those studies also either used fictitious COVID-19 vaccines^17,18^ or did not name specific approved COVID-19 vaccines^19^. As such, participants either had limited knowledge of, or investment in, the framed vaccines. As the pandemic has progressed, media discourse^20,21^ combined with direct and socially-observed experience of COVID-19 side effects^22^, means it is essential to understand whether positive framing is effective for real-world vaccines where prior knowledge and experience exists. This is particularly important because there is reason to believe that prior knowledge regarding a given COVID-19 vaccine may moderate the strength of any positive framing effect.

Research has shown that greater relevance or belief in a treatment or issue decreases the efficacy of different forms of framing^23,24^, including positive framing on perceptions of hypothetical vaccines^23,25^. The effect of positive framing on vaccine intention may therefore be limited to less familiar vaccines. Even if so, positive framing may still hold utility. New composition changes to COVID-19 booster vaccines have been recommended^26^ and are currently being developed^27^ to protect against emerging variants. Further, many booster programmes (e.g., in the United Kingdom) require switches from an experienced vaccine (e.g., AstraZeneca Vaxzevria) to a less familiar one (e.g., Moderna Spikevax). Positive framing may therefore be beneficial for increasing uptake for novel vaccines and switches to less familiar vaccines. Yet, because research on positive framing has largely focused on fictitious medications and patient scenarios^23,25,28-32^, the extent to which familiarity moderates the effect of positive framing is currently unclear. Therefore, to understand the extent to which positive framing could be deployed to improve global vaccine uptake to combat COVID-19, it is critical to test the efficacy of positive framing for genuine familiar and unfamiliar vaccines.

In this pre-registered study (aspredicted.org/53ph4.pdf), positive attribute framing was applied to side effects from genuine manufacturer Patient Information Leaflets (PILs) for the AstraZeneca, Pfizer, and Moderna vaccines and compared to standard negative wording. Participants were randomised to read positive or negatively framed PILs for the same vaccine they had previously received (either AstraZeneca/Pfizer), a familiar vaccine in the UK context (Pfizer/AstraZeneca), or an unfamiliar vaccine (Moderna). It was hypothesised that a) positive framing would increase booster intention, and b) the effect of positive framing would decrease with vaccine familiarity. Following previous research^14,28,33-35^ secondary outcome variables concerning booster side effect severity, perceived risk, and booster acceptance, as well as prevalence judgments, were explored as potential mediators of the framing effect on vaccine intention (see Supplementary Materials 1).

## Results

### Overview

Participants (*N*=1,222) were recruited from the United Kingdom via Pureprofile, an international recruitment service, between 27^th^ October and 8^th^ November 2021. All were pre-screened (see Methods) with data removed on pre-registered quality control criteria (see Supplementary Materials 2). Only those who had received two doses of either the AstraZeneca or Pfizer COVID-19 vaccines were eligible to participate, with participants stratified by their previous vaccine type.

Participants from each stratum were subsequently randomised to one of six experimental conditions (2(Framing)*3(Familiarity) factorial design; see Figure 1). Baseline ratings were collected regarding primary and secondary outcome measures—all recorded on a 100-point VAS. The primary outcome was the participant’s intention to receive a booster vaccine. Secondary outcomes were: 1) perceived booster side effect severity; 2) perceived booster risk; 3) booster acceptance. Wording of items can be found in Figure 1. Familiarity with the side effects of the AstraZeneca, Pfizer, and Moderna vaccines were additionally assessed (100-point VAS) to determine whether side effect knowledge corresponded with the predetermined factorial categories of vaccine Familiarity (i.e., Same > Familiar > Unfamiliar).

**Figure 1:**
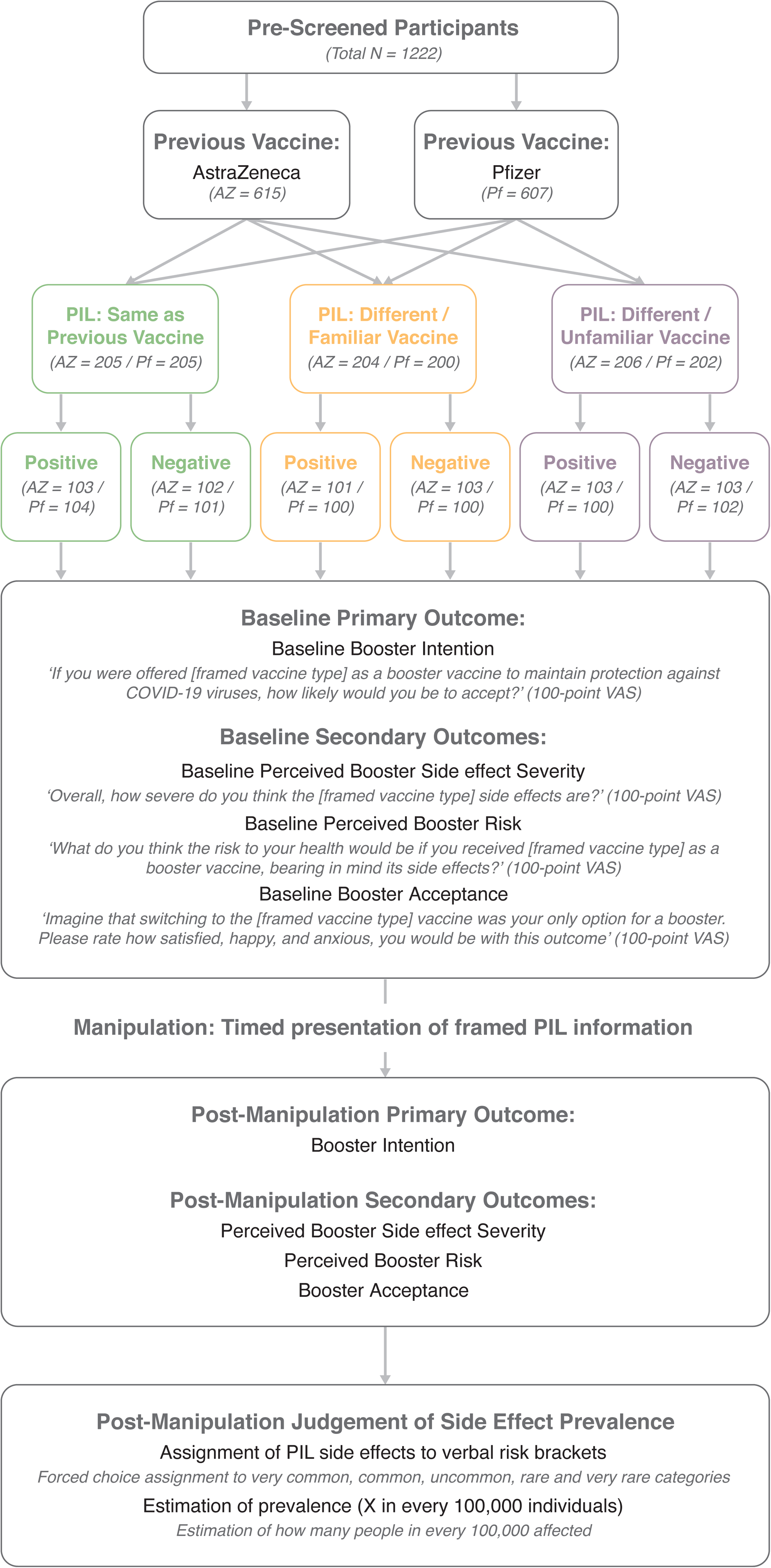
Overview of the experimental design, including the item wording for primary and secondary outcomes (*Nb*. analyses concerning secondary outcomes are contained in Supplementary Materials 1).

Framed side effect information was subsequently presented dependent on condition using genuine manufacturer PILs for the AstraZeneca, Pfizer, and Moderna vaccines, with side effects presented in standard EU verbal prevalence categories and frequency bands. Positive framing was applied to the frequency bands (for example see Figure 2; full wording and PILs presented as Supplementary Materials 3-4). To manipulate familiarity, participants were randomised to view PILs from the following conditions: ‘Same’ (PIL for the COVID-19 vaccine previously received: AstraZeneca-AstraZeneca|Pfizer-Pfizer); ‘Familiar’ (PIL for a common vaccine not previously received: Pfizer-AstraZeneca|AstraZeneca-Pfizer); ‘Unfamiliar’ (PIL for a less common vaccine in the UK: AstraZeneca-Moderna|Pfizer-Moderna). Familiarity was judged on UK data (22^nd^ September 2021), where fewer Modena second doses (1.2 million) had been administered relative to the Pfizer and AstraZenca vaccine (19.4 and 24.0 million respectively)^36^. After timed PIL presentation of 2 minutes, all participants provided post-manipulation ratings for the primary and secondary outcomes (as above). Participants were then presented with 14 side effects from their assigned PIL and were required to: 1) assign each to a verbal prevalence category (i.e., very common, common, uncommon, rare, very rare); 2) estimate frequency of occurrence (number of people in 100,000 expected to experience the side effect (results in Supplementary Materials 1.4.1).

**Figure 2:**
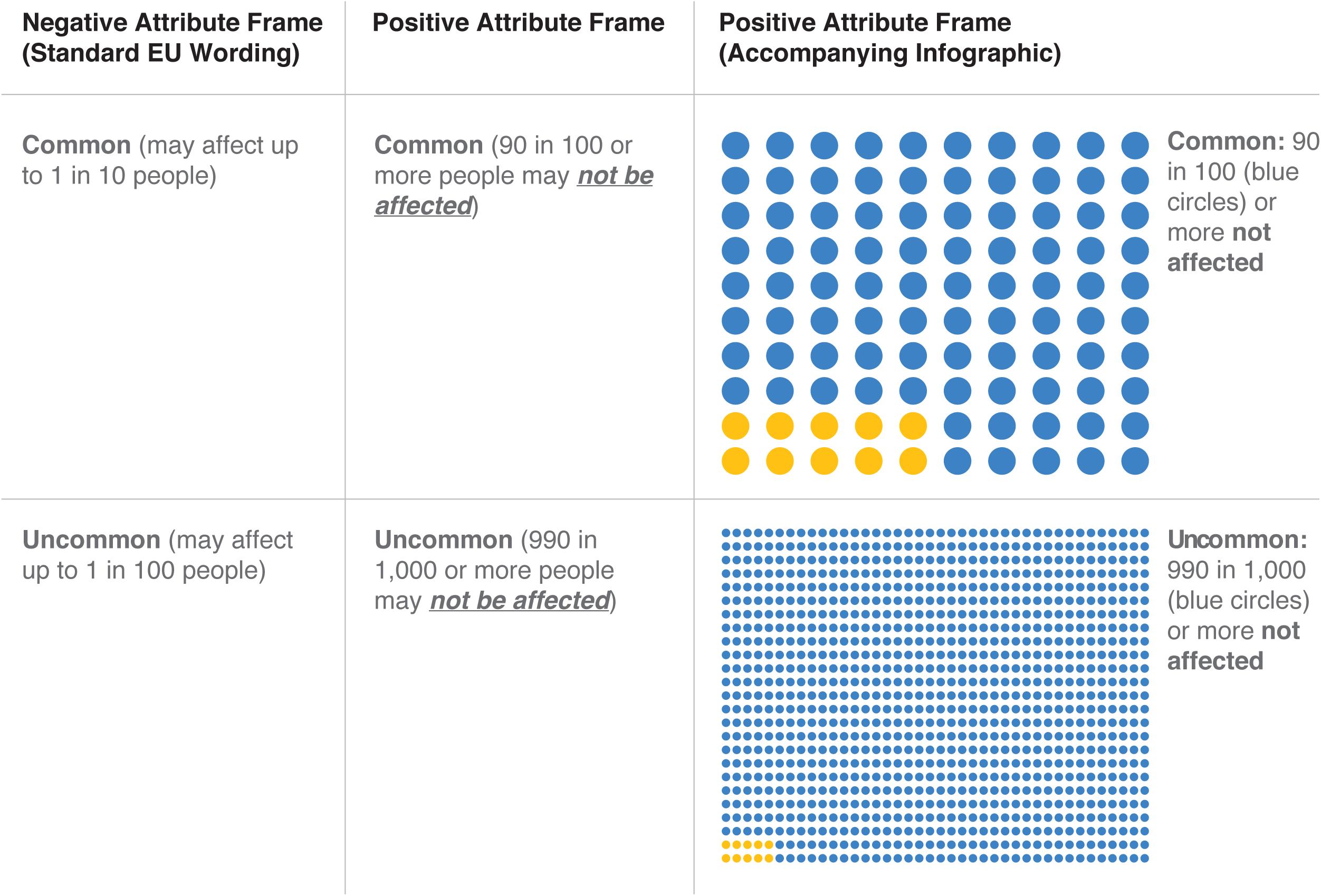
Positive and negative wording used to frame common and uncommon side effects (wording for all prevalence categories can be found in Supplementary Materials 3)

### Descriptive Statistics

#### Sample demographics

Information regarding participant location, age, education, and employment can be found in Figure 2a-b. Participants were 52.5 years of age on average (range=18-95). They resided across most postal areas in the UK, with the largest proportion from London district (*N*= 53), Birmingham (*N*=35), and Belfast (*N*=34). Only Harrogate, and the Orkney and Shetland Islands were not represented. The largest proportion of respondents identified as British (*N*=1,064; 87%), female (*N*=706; 58%), had completed undergraduate-level education (*N*=413; 34%), and were in full-time employment (*N*=407; 33%). Descriptive statistics for the full sample, and by condition, are presented as Supplementary Materials 5-6.

#### COVID-19 exposure and vaccine history

Most participants (*N*=1,074; 88%) had not personally been infected with COVID-19, while 565 (46%) reported infections among family and close friends. On average, participants had their last COVID-19 vaccine 4.8 months ago. The largest proportion reported not experiencing side effects to either dose (*N*=579; 47%). Of those who did, the first dose was most associated with severe side effects (*N*=372; 30%).

### Primary Analysis

#### Knowledge of vaccine side effects mirrors categorical levels of the Familiarity factor

To determine whether side effect familiarity corresponded with the predetermined factorial categories of Familiarity (i.e., Unfamiliar, Familiar, Same), a within-subjects one-way ANOVA (with Greenhouse–Geisser correction) was run on side effect familiarity ratings collected for the three vaccine types prior to the experimental manipulation. A robust main effect of Familiarity was observed (*F*(1.86, 2272.18)=659.17, *p*<.001, *η*_*p*_^*2*^=.35). Awareness of side effects increased with Familiarity; being higher for the Same vs. Familiar vaccine (*t*(1221)=14.11, *p*<.001, Cohen’s *d*_*z*_ =.40), and for the Familiar vs. Unfamiliar vaccine (*t*(1221)=23.97, *p*<.001, Cohen’s *d*_*z*_ =.69). Mean differences are presented in Figure 4e.

**Figure 3:**
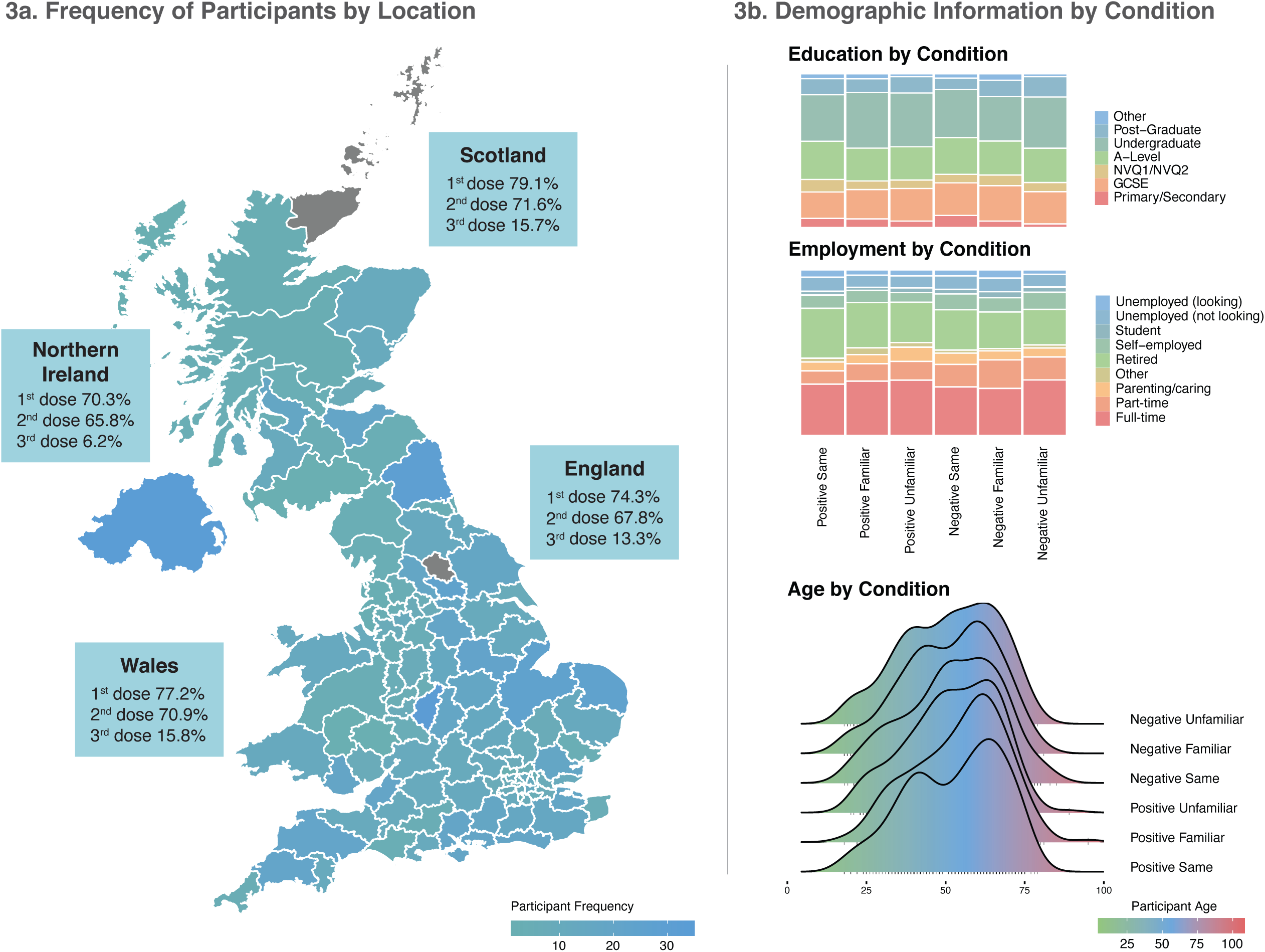
Demographic Information, where 3a plots the frequency of participants from each postal area of the UK against the vaccination rates reported by the UK government on the 3^rd^ November (the final week of data collection) and 3b depicts education and employment categories, and age range, across experimental conditions. *Nb*. 6 participants, 0.5% of the sample, are not represented in 3a due to not providing a valid postal code.

**Figure 4:**
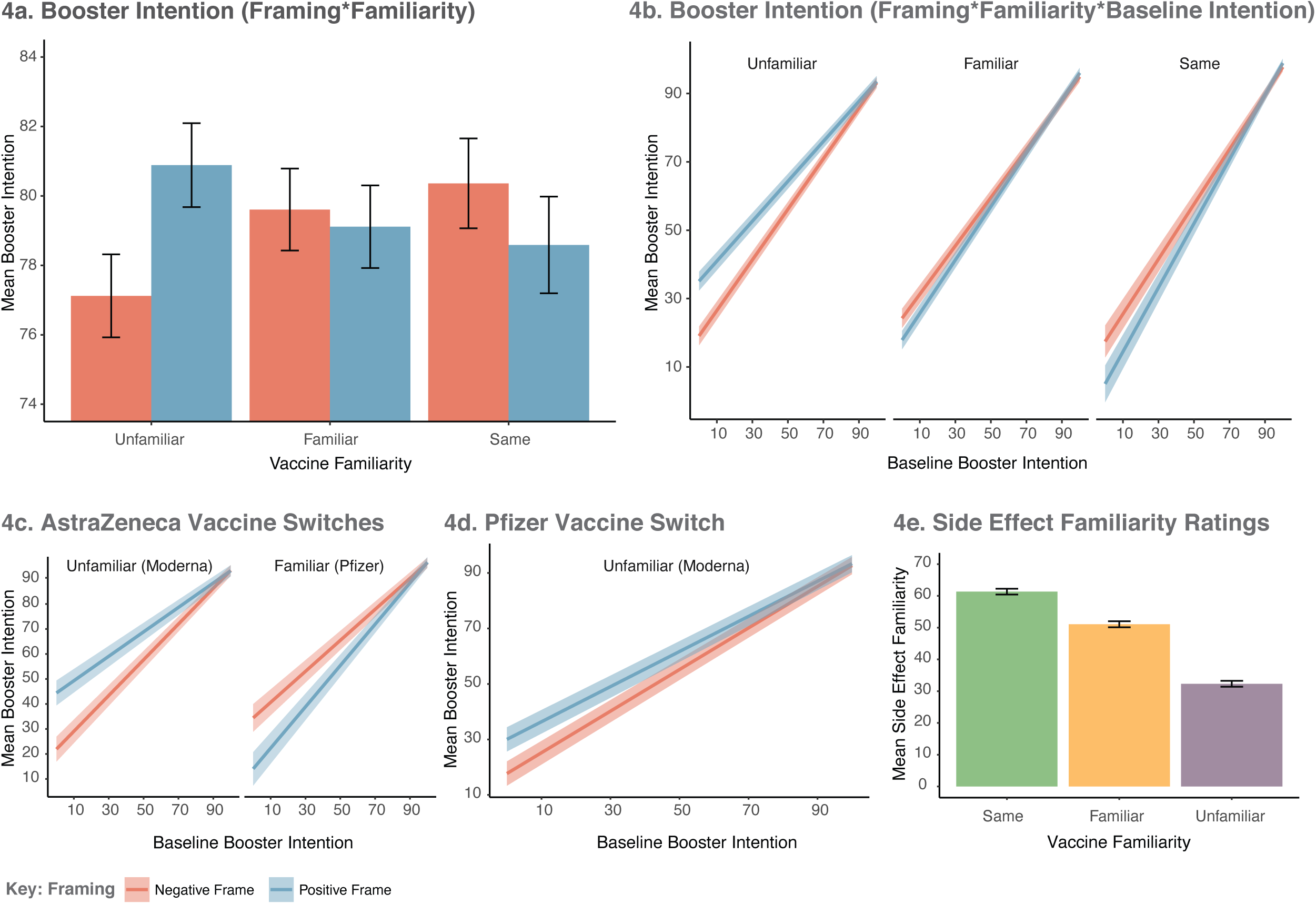
Model estimated mean differences in the primary outcome (Booster Intention), depicted for the whole sample (4a and 4b), and for realistic switches occurring as part of the UK booster programme (4c and 4d). 4e presents data demonstrating that side effect familiarity ratings scaled with the factorial levels of Vaccine Familiarity. All error bars represent ± 1SEM.

#### Baseline vaccine intention

A 2(Framing)*3(Familiarity) ANOVA was conducted on Baseline Booster Intention scores to assess the presence of between-group differences. Baseline Booster Intention was anticipated to be high across conditions (see pre-registration), which was confirmed in the present sample (*M*=78.36 (/100-point VAS), *SD*=31.65; range:0-100). However, an unanticipated significant effect of Familiarity was observed(*F*(2, 1216)=49.51, *p*<.0001, *η*_*p*_^*2*^=.075). This effect reached significance for the orthogonal contrast comparing the Same vs. Other (i.e., combined Familiar and Unfamiliar) vaccine types (*F*(1, 1216)=96.51, *p*<.0001, *η*_*p*_^*2*^=.074), but not for the Familiar vs. Unfamiliar comparison (*F*(1, 1216)=2.42, *p*=.120, *η*_*p*_^*2*^=.002), indicating higher intentions for previously experienced vaccines.

#### Effect of Framing and Familiarity on the intention to be vaccinated

The pre-registered primary analysis plan was a 2(Framing: Positive vs. Negative)*3 (Familiarity: Same, Familiar, Unfamiliar) factorial ANCOVA, with Baseline Booster Intention for the vaccine presented in the participant’s assigned PIL as the covariate, and Post-Manipulation Booster Intention the outcome. However, the fact that Baseline Intention unexpectedly differed systematically with Familiarity meant that including it as a covariate without including its interactions with the manipulated variables (Framing and Familiarity) would violate the assumptions of ANCOVA^37^. To address this, we extended the model to include the interactions between Baseline Booster Intention and Framing and Familiarity in all analyses, as recommended elsewhere^38^. Pre-specified orthogonal contrasts for Familiarity were: Contrast1 (Same vs. Other [Familiar and Unfamiliar combined]); Contrast2 (Familiar vs. Unfamiliar). A Framing*Familiarity interaction was hypothesised, with the effect of Framing (increased Booster Intention following Positive vs. Negative framing) reducing with increased Familiarity.

Main effects analysis revealed the anticipated Framing*Familiarity interaction (*F*(2, 1210)=10.75, *p*<.0001, *η*_*p*_^*2*^=.018). Specifically, Framing interacted with Contrast1 (Same vs. Other: *F*(1, 1210)=5.07, *p*=.025, *η*_*p*_^*2*^=.004) and Contrast2 (Familiar vs. Unfamiliar: *F*(1, 1210)=16.46, *p*=.0001, *η*_*p*_^*2*^=.013). As demonstrated in Figure 4a, this pattern of results was driven by Positive Framing increasing Booster Intention for the Unfamiliar vaccine. However, this was superseded by a three-way interaction with Baseline Booster Intention (*F*(2, 1210)=7.65, *p*=.0005, *η*_*p*_^*2*^=.013), represented at both Contrasts (Baseline*Framing*Contrast1: *F*(1, 1210)=4.39, *p*=.036, *η*_*p*_^*2*^=.004 | Baseline*Framing*Contrast2: *F*(1, 1210)=11.19, *p*=.0008, *η*_*p*_^*2*^=.009). Figure 4b depicts this interaction. Positive Framing had limited efficacy at high levels of Baseline Intention across Conditions but took effect for the Unfamiliar Vaccine when model estimated Baseline Intention scores were lower than ∼80/100. At very low levels of Baseline Intention (i.e., VAS=0), model predicted Booster Intention Post-Manipulation increased from *M*=19.09 (*SEM*=2.76, 95% CIs[13.68, 24.50]) for the Negative Frame, to *M*=35.11 (*SEM*=2.77, 95% CIs[29.68, 40.50]) for the Positive Frame. Full model output is included as Supplementary Materials 7. We note that the same interaction between Framing and Familiarity was observed in the planned but invalid model excluding the interaction between these factors and Baseline Booster Intention (see Supplementary Materials 8).

#### The effect of Framing on vaccine switches

Analyses were performed to investigate the effect of framing on pre-registered and realistic vaccine switches occurring as part of the UK’s booster programme. Specifically, those without medical exemption, who previously received the AstraZeneca vaccine will be required to switch to Pfizer or Moderna, while those who received Pfizer may be required to switch to Moderna.

First, previous vaccine type (AstraZeneca/Pfizer) was entered as a factor in the ANCOVA model above to check for interactions with Framing, Familiarity, and Baseline Booster Intention. While there was a main effect of previous vaccine type (*F*(1, 1198)=6.36, *p*=.012, *η*_*p*_^*2*^=.005), with those receiving AstraZeneca reporting increased Booster Intention (*M*=80.83, *SEM*= 0.74, 95% CIs [79.37, 82.29]) compared to Pfizer (*M*=78.01, *SEM*= 0.74, 95% CIs [76.57, 79.46]), there were no two- or three-way interactions with Framing or Familiarity (all *ps*>.05; see Supplementary Materials 9).

### Previous vaccine: AstraZeneca

Representative of a switch from AstraZeneca to Pfizer or Moderna, a 2(Framing)*2(Familiarity: Familiar (Pfizer), Unfamiliar (Moderna)) ANCOVA (modelling interactions between covariate and factors), was conducted among those who had previously received the AstraZeneca vaccine (*N*=410). A three-way Framing*Familiarity*Baseline Intention interaction was observed (*F*(1, 402)=11.38, *p*=.0008, *η*_*p*_^*2*^=.028). As demonstrated in Figure 4c, in the case of the Unfamiliar vaccine (Moderna) Booster Intention was increased in the Positive Frame at low levels of Baseline Booster Intention. However, the inverse of this pattern was observed for the Familiar vaccine (Pfizer), where Positive Framing decreased Booster Intention at low levels of Baseline Intention.

### Previous vaccine: Pfizer

Representative of a switch from Pfizer to Moderna, a one-way ANCOVA investigating the effect of framing on the perception of the Unfamiliar (i.e., Moderna) PIL was performed on the data of those who had previously received the Pfizer vaccine (*N*=202). There was a main effect of Framing (*F*(1, 198)=3.98, *p*=.048, *η*_*p*_^*2*^=.020), but no statistically significant Baseline Booster Intention*Framing interaction (*F*(1, 198)=1.80, *p*=.181, *η*_*p*_^*2*^=.009). However, as demonstrated in Figure 4d, slopes for the Positive and Negative Frame converged at high levels of Baseline Booster Intention.

## Discussion

Message framing has been suggested as a potential intervention to increase COVID-19 vaccine uptake^12^. We examined the effect of positive and negative attribute framing of side effect information on booster intentions for three genuine COVID-19 vaccines varying in familiarity.

Positive framing successfully increased booster intention for the unfamiliar vaccine (i.e., Moderna), but reduced intention for the vaccine previously received, as well as for a switch to Pfizer among those previously receiving AstraZeneca. In all cases, effects were strongest at low baseline booster intentions. Increasing booster acceptance among those with low intentions is of substantial importance in protecting against infection from, and transmission of, COVID-19 viruses. Critically, our data suggest that any intervention intending to employ attribute framing should be carefully tailored to match the framed information (positive vs. negative wording) with vaccine familiarity. Specifically, positive framing appears to have significant potential in situations where a novel vaccine or composition changes are being introduced^26,27^. By contrast, positive framing may actually be harmful when the vaccine is familiar.

The effect of positive attribute framing on booster intentions for the Unfamiliar vaccine is consistent with medical decision-making research. In these studies, framed information has typically been presented regarding fictitious medications and patient scenarios^23,25,28-32^. When employing real treatments, data has been collected from samples where participants were completely^15,39^ or largely^40^ naïve to the framed treatment, or where prior treatment experience was not assessed^16,41^. The current data thereby provide new insights into the effect of framing.

Under conditions directly relevant to the COVID-19 pandemic (i.e., for real vaccines, at high levels of public involvement), the benefit of positive attribute framing was found to wane, or be reversed, as familiarity and prior experience with the framed vaccine increased. As such, calls for all PILs to employ positive framing as standard (e.g.,^28,42^) appear premature. Instead, negative framing, the standard form for communicating side effect information within the European Union, appears beneficial when treatments are well known.

The reduced efficacy of positive framing with increased vaccine familiarity could be explained by a current theory of attribute framing that posits an interaction between familiarity (a manifestation of psychological distance) and the valence of the message surrounding a given attribute or event (e.g., the experience of vaccine side effects). At closer psychological distances (e.g., for vaccines that are more familiar and more likely to be received), negatively framed information has been shown to be more persuasive^43^. Further experimental research is needed to test this theory, while considering alternative explanations, such as the role of potential backfire effects in persuasive or corrective messaging. Such effects are known to impact attitudes surrounding the COVID-19 pandemic^44^ and have been demonstrated to lower intentions for other vaccine types at high levels of concern^45^. However, when assessed in conjunction, current results highlight the fact that any intervention that strives to apply positive framing across all vaccine types, irrespective of familiarity, should be treated with caution.

The psychological mechanisms underlying the effect of positive framing on booster intentions remain unclear. We measured secondary variables as potential mediators. However, results did not mirror those obtained for the primary outcome – booster intention (see Supplementary Materials 1). While changes in secondary variables (side effect severity and booster acceptance) were observed with framing, post-hoc analysis (see Supplementary Materials 10) plotting the Familiarity*Framing*Baseline interaction for those who had high vs. low baseline booster intent, suggested that these framing-induced changes largely occurred among those with high vaccine intention at baseline. As these participants also showed limited effect of framing on their behavioural intention to be vaccinated, the relationship between booster intention and our secondary predictors appears orthogonal. Investigation of other factors combined with qualitative research may be better positioned to identify the driving factors behind the effect of framing on COVID-19 booster intentions. Further, we note that, consistent with previous reports^35,46-49^, prevalence judgements were poor (<∼35% accuracy; Supplementary Materials 1.4). This appeared exacerbated among those receiving positive framing, but again did not differ by familiarity. As side effects differed by PIL, the current study was designed only to test for general inaccuracies in side effect representation and not systematic over- or under-estimation. Experimental studies are therefore needed to assess precisely how any inaccuracies associated with positively framed COVID-19 vaccine information manifest.

The primary strength of the present study is the application of attribute framing to real COVID-19 vaccine information. The PILs employed here are displayed on government and NHS websites in the UK, forming a primary official source of information regarding COVID-19 vaccination. Our findings therefore have real-world implications, demonstrating that the wording of PILs can directly impact the intention to receive a booster vaccine among individuals for whom this decision is both directly relevant and imminent. There are of course some limitations worth noting including the collection of cross-sectional data that limits an assessment of the durability of the framing effect, as well as a sample located within a single country. Given global differences in booster policy, cross-cultural replication of results is required to ensure results are not contextually limited to the UK. While vaccine intention has been demonstrated to be a strong predictor of vaccine uptake (e.g.,^50-52^), including for COVID-19 vaccination^53^, we do not assume that the two are synonymous^54^. While beyond the scope of the present study, we recommend that future research incorporate longitudinal designs where the rate of conversion from intention to vaccine uptake can be tracked. Further, present results are specific to booster intentions among those already vaccinated. While side effect apprehension has been associated with hesitancy regarding COVID-19 vaccination^9^ and booster vaccination^11^, whether a similar pattern of results would hold among those who have never been vaccinated is unknown.

In summary, the present study demonstrates that the ability of positive framing to successfully increase booster intention for genuine COVID-19 vaccines is critically moderated by the familiarity of that vaccine. Positive framing can improve vaccine intention for unfamiliar vaccines, but may actually decrease intentions for familiar vaccines. The data therefore provide novel insights into the benefits of positive framing for COVID-19 vaccines and beyond. As such, we recommend that if positive attribute framing is to be employed, close attention must be paid to the type of treatment being framed as well as the likely recipients of the framed information.

Importantly, in the context of the current COVID-19 pandemic, positive framing appears capable of improving uptake of COVID-19 vaccines when switches or new developments require individuals to receive unfamiliar vaccines, as is the case for many booster vaccine programmes globally.

## Methods

### Recruitment and Sample

Recruitment took place in the initial weeks of the UK booster programme (27^th^ October -8^th^ November 2021). Figure 3a provides a summary of the number of boosters administered in the UK at the time of data collection. The sample was obtained from Pureprofile, an ISO-certified panel provider for online research, and balanced on national quotas for age, gender, and region. All potential participants were screened using the following inclusion criteria: 18 years of age or older, currently residing in the UK, self-reported English fluency, previously received two doses of either the Pfizer or the AstraZeneca COVID-19 vaccines (no other combination), not having received a third COVID-19 booster vaccine, and no known medical reason (e.g., allergy) prohibiting potential administration of the Pfizer, AstraZeneca, or Moderna COVID-19 vaccines. After screening, 1896 participants provided electronic consent and 1459 completed the experiment. To reduce statistical noise due to random and inconsistent responding, 237 completing participants were removed based on pre-registered quality control criteria (see Supplementary Materials 2). The experiment was approved by the University of Sydney Human Research Ethics Committee. All completing participants were provided with an electronic debrief at the end of the survey and were paid £3.50 for a ∼15-minute survey.

### Data Collection and Procedure

Cross-sectional data were collected online via Qualtrics, with the survey accessible to personal computer, tablet, and smartphone. The ‘force response’ option was used to ensure complete cases for all outcome variables. Participants completed the survey in one sitting and were not able to return to the study URL. After pre-screening, those meeting the inclusion criteria were provided with a participant information statement detailing the aims of the study and gave their digital informed consent. Those not meeting these criteria were directed away from the survey.

Participants completed demographic items and identified which COVID-19 vaccine they had previously received (AstraZeneca or Pfizer). Stratified randomisation to the six experimental groups occurred at this point using the inbuilt Qualtrics randomisation function. Quotas were set to limit data collection to 600 participants from each vaccine type (AstraZeneca/Pfizer), with 100 from each group randomised to the six experimental conditions. Because Qualtrics tallies quotas on survey completion but does not account for participants currently in the experimental pipeline, the final sample contained 22 more participants than projected. Statistical analysis did not take place until after exclusions had been made and all quotas were closed.

After randomisation, participants responded to items concerning the number of months since their last COVID-19 vaccine and familiarity with side effects of the three framed vaccines. For each vaccine type, participants then rated their booster intention, perceived booster side effect severity, perceived booster risk, and booster acceptance (satisfaction, happiness, and anxiety). Responses made to the vaccine type that matched with the experimental condition to which the participant had been assigned were employed as baseline measures. Responses to all other vaccine types were recorded for use in a concurrent, but separate, pre-registered study (#78370; aspredicted.org/8e6af.pdf). PILs containing the positively or negatively framed side effect information were then displayed for two minutes, using a timer embedded in the survey. Participants could not proceed until this time had elapsed. Post-manipulation booster intention, perceived booster side effect severity, perceived booster risk, and booster acceptance were then recorded. Finally, fourteen side effects described in the PILs were presented, with participants required to assign each to a verbal prevalence descriptor (very common, common, uncommon, rare, very rare) and estimate how many people (out of 100,000) taking the vaccine would experience the side effect. On completing the survey, all participants were provided with an electronic debrief for download and URLs to the UK government landing page where the original PILs for the vaccines used in the study could be found.

Several additional items concerning general COVID-19 booster intentions, perceived risk of previous COVID-19 vaccines, specific COVD-19 vaccination side effects, general perceptions of COVID-19 and COVID-19 vaccinations, and severity of previous COVID-19 infection, were included in the survey for use in a separate pre-registered study (for information see: #78370; aspredicted.org/8e6af.pdf).

### Survey Materials

#### Demographic information

Participants responded to items concerning their age, gender, ethnicity, highest level of education and employment status, and geographic region (postal area code). Exact wording can be found in the Demographic Information section, Supplementary Materials 5.

#### Previous exposure to COVID-19

Items were employed to capture personal exposure to COVID-19, as well as exposure through close friends and family. Item wording (*To your knowledge, are you, or have you been, infected with COVID-19? / To your knowledge, have any of your close family members or friends been infected with COVID-19?)* was taken from the WHO BeSD documentation^12^.

#### Previous COVID-19 vaccination history

Previous COVID-19 vaccine (Pfizer/AstraZeneca) was recorded as a forced-choice option. Participants indicated the number of months since their last COVID-19 vaccine, and whether their most severe side effects occurred to their first dose, second dose, whether they were equal across doses, or not experienced at all.

#### Familiarity with COVID-19 vaccine side effects

For the three framed vaccines, participants were asked to rate their “*familiarity with the potential side effects*” on a 100-point VAS, with anchor labels (*‘not at all familiar’* / *‘extremely familiar’*) positioned to the left and right of the scale. Those who had not heard of the vaccine were asked to check a separate ‘*not heard of vaccine’* box but received a score of zero (*not at all familiar*). This response-type was used to exclude inconsistent responders (see Supplementary Materials 2).

#### Baseline measures: COVID-19 vaccine perceptions

Four items were used as baseline measures. All were rated on a 100-point VAS. For each of the three vaccines, participants responded to questions concerning: 1) booster intention *‘If you were offered* [vaccine type] *as a booster vaccine to maintain protection against COVID-19 viruses, how likely would you be to accept?’* (labels: *‘definitely would not accept vaccine’* vs. *‘definitely would accept vaccine’*); 2) perceived booster side effect severity *‘Overall, how severe do you think the* [vaccine type] *side effects are?’* (labels: *‘not at all severe’* vs. *‘extremely severe’*); 3) perceived booster risk *‘What do you think the risk to your health would be if you received* [vaccine type] *as a booster vaccine, bearing in mind its side effects?’* (labels: *‘extremely low risk’* vs. *‘extremely high risk’*); 4) booster switch perceptions *‘Imagine that switching to the* [vaccine type] *vaccine was your only option for a booster. Please rate how satisfied, happy, and anxious, you would be with this outcome’* (labels: *‘not at all’* vs. *‘extremely’*). Satisfaction, happiness, and anxiety were rated as sub-items, each yielding a score of 0 – 100. Where the vaccine type was the same as that previously received by the participant, wording was changed from ‘switching to’ to ‘continuing with’. Wording of 1 – 3 was adapted from previous research^35^.

#### Patient Information Leaflets (PILs): Experimental manipulation

Genuine PILs for the AstraZeneca, Pfizer, and Moderna vaccine were abridged to include the manufacturer’s description of each vaccine, what it is used for, and critically, the possible side effects resulting from administration. Side effects were retained in their original form and order. For both the negatively and positively framed PILs, verbal descriptors were in keeping with those published by the manufacturer (i.e., very common, common, uncommon, rare, very rare, not known). Wording of assigned frequency bands employed in the negatively framed PILs was also identical to that of the manufacturer. However, this was inverted in the positively framed PILs to stress the number of individuals *not* affected (e.g., “Common (90 in 100 or more people may *<u>not be affected</u>*)”). As multi-modal forms of side effect presentation (e.g., written, pictorial, verbal) may elicit larger framing effects^14^, and numeracy is less likely to moderate the effect of attribute framing for graphical presentations^55^, positive PILs additionally included a graphical representation of side effect risk to enhance the manipulation. For an example of the wording used to frame side effects see Figure 2.

#### Post-manipulation measures: COVID-19 vaccine perceptions

Participants responded to the four vaccine perception items listed above under Baseline Measures (booster intention, booster side effect severity, booster risk, booster acceptance), but only in relation to the vaccine type outlined in the PIL to which they were assigned.

#### Post-manipulation side effect perceptions

To determine whether positive framing augments the perception of side effect occurrence relative to true prevalence rates, fourteen side effects were presented from each PIL. Eleven of these were associated with concrete prevalence brackets in the original PILs, while the remaining three were assigned to the ‘unknown’ category (i.e., could not be estimated at the time of PIL publication). For each side effect, participants were required to identify the correct verbal descriptor (very common, common, uncommon, rare, very rare): “*based on the information that you read, how common do you think* [side effect] *is?”* (forced-choice answer). They were then asked, “*In 100,000 people, how many do you think would experience* [side effect] *if they received a* [framed vaccine name] *booster vaccine?”* (free-response, limited to numbers at up to 10 decimal places). The three ‘unknown’ side effects were analysed separately to determine the effect of framing when prevalence rates are not provided.

### Statistical Analysis

Statistical analysis was performed using R (version 4.1.1). Statistical analyses were a combination of ANCOVAs (where baseline measures were available) or ANOVAs, reported with Type III Sum of Squares. An alpha of .05 was accepted as the threshold for statistical significance. Sample size (estimated *N*=1200) was calculated based on an *a priori* power analysis (95% power, alpha=.05, effect size *f*^*2*^=0.02) for a separate study run concurrently that contained more predictor variables (*N*=9), and therefore required more power, than the study presented here (further details contained in the study pre-registration form). An effect size for attribute framing was additionally derived from previous research (average effect size *r*= 0.175)^14^. An *a priori* power analysis based on this effect size required a total of 491 participants, providing reassurance that the projected sample size for the study above provided ample power to detect an effect of framing in the present study.

## Data Availability

All data produced are available online through the Open Science Framework repository

https://osf.io/d5cvn/?view_only=cb90eca9ddf446c8901652ab0344fa05

## Sources of Funding

This research was supported by Australian Research Council grants DP180102061 and DP200101748. The funding body had no involvement in study design, analysis, interpretation, writing, or the decision to submit the present article for publication.

## Data Availability

The code and raw data necessary to replicate the reported analysis is available through the Open Science Framework repository (means/SEMs needed to reproduce the analysis figures are included with the raw data): https://osf.io/d5cvn/?view_only=500c83d90bb9416796e464108ad2fc41

## Author contributions

KB and BC conceived the experimental design. KB was responsible for collecting and analysing the data. KB wrote the first draft of the article. BC edited and contributed to the final version.

## Notes

### Competing Interest Statement

The authors have declared no competing interest.

### Clinical Protocols

https://aspredicted.org/53ph4.pdf

### Author Declarations

University of Sydney Human Research Ethics Committee of the University of Sydney gave ethical approval for this work

### Summary of Updates

Secondary analyses have been moved to Supplementary Materials

