## Supplementary Materials for "Positive attribute framing increases COVID-19 booster vaccine intention for unfamiliar vaccines"

#### Supplementary Materials 1: Analysis of Secondary Variables

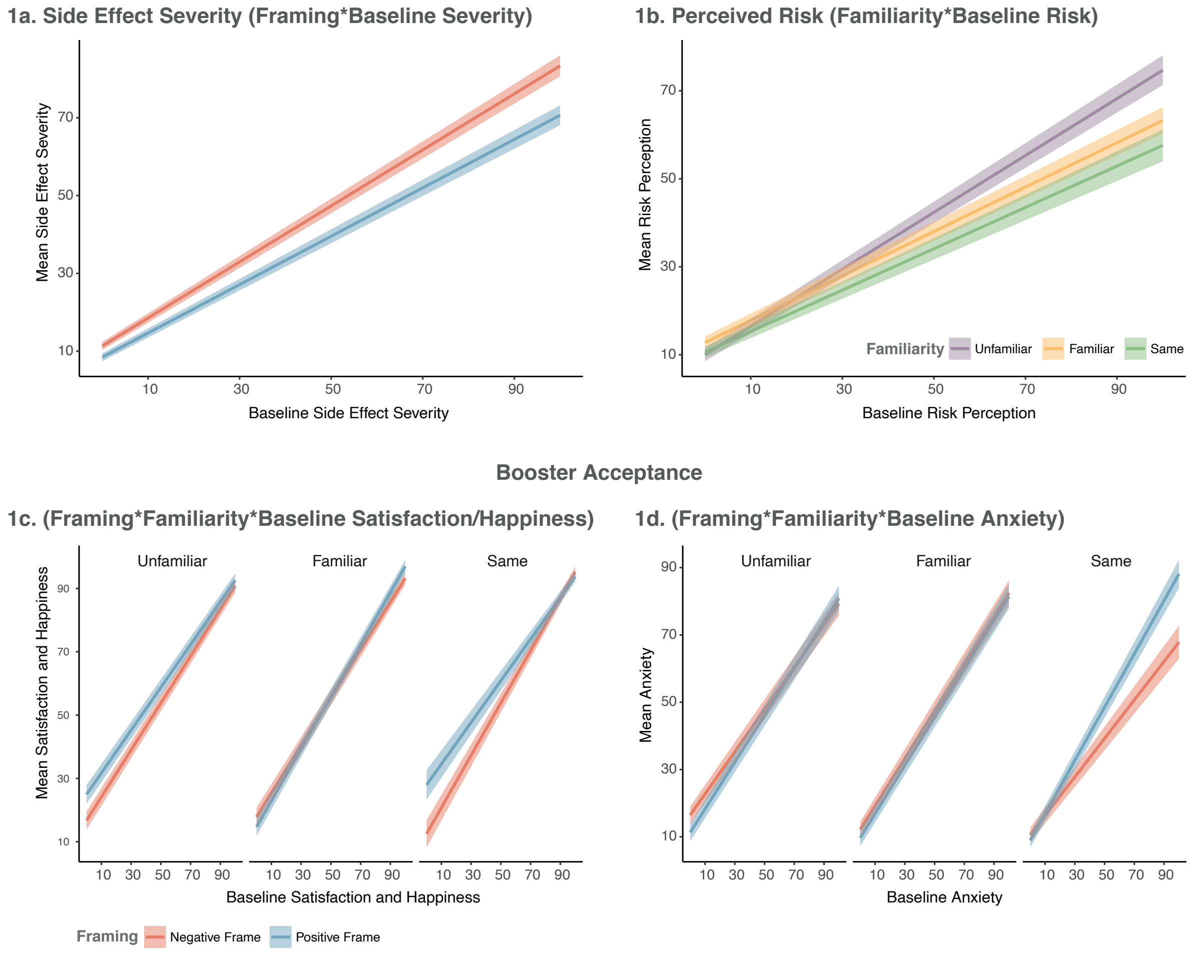


Supplementary Figure 1: Mean values for secondary outcome measures, where 1a depicts model-estimated mean differences regarding side effect severity ratings, 1b perceptions of vaccine risk, 1c combined satisfaction and happiness associated with the framed vaccine, and 1d anxiety surrounding the framed vaccine. All error bars represent +/- 1SEM.

#### Supplementary Materials 1.1: Booster Side Effect Severity

##### Summary

Perceived side effect severity associated with the vaccines was measured pre- and post-manipulation. A 2(Framing) * 3(Familiarity) ANCOVA, including interactions with the covariate (Baseline Side Effect Severity), revealed a main effect of Framing ($F(1, 1210)=3.92$, $p=.048$, $np2=.003$), and a Baseline Severity by Framing interaction ($F(1, 1210)=4.52$, $p=.034$, $np2=.004$). There were no other two- or three-way interactions (all $ps>.35$).

As depicted in Supplementary Figure 1a, reading about side effects in any frame increased the perception of their severity (i.e., post-manipulation model estimated means are above zero, for all levels of the baseline covariate). However, Positive Framing resulted in a lower perception of side effect severity, which was strongest when side effects were perceived as severe at baseline.

##### Booster Side Effect Severity: Statistical Models

- sesev_fv_t2 (perceived side effect severity associated with the framed vaccine post-manipulation)
- sesev_fv_t1 (perceived side effect severity associated with the framed vaccine at baseline)

##### Main Effects Model

| | Sum Sq | Df | F value | Pr(>F) |
|---|---|---|---|---|
| (Intercept) | 57189.5779 | 1 | 183.2956 | 0.0000 |
| sesev_fv_t1 | 263557.1698 | 1 | 844.7145 | 0.0000 |
| Framing | 1222.8893 | 1 | 3.9194 | 0.0480 |
| Familiarity | 40.5057 | 2 | 0.0649 | 0.9372 |
| sesev_fv_t1:Framing | 1411.1856 | 1 | 4.5229 | 0.0336 |
| sesev_fv_t1:Familiarity | 654.8262 | 2 | 1.0494 | 0.3505 |
| Framing:Familiarity | 157.2217 | 2 | 0.2520 | 0.7773 |
| sesev_fv_t1:Framing:Familiarity | 314.8565 | 2 | 0.5046 | 0.6039 |
| Residuals | 377528.9467 | 1210 | NA | NA |

| | eta.sq | eta.sq.part |
|---|---|---|
| sesev_fv_t1 | 0.3935 | 0.4111 |
| Framing | 0.0018 | 0.0032 |
| Familiarity | 0.0001 | 0.0001 |
| sesev_fv_t1:Framing | 0.0021 | 0.0037 |
| sesev_fv_t1:Familiarity | 0.0010 | 0.0017 |
| Framing:Familiarity | 0.0002 | 0.0004 |
| sesev_fv_t1:Framing:Familiarity | 0.0005 | 0.0008 |

**Model with pre-registered contrasts for Vaccine Familiarity**

| | Sum Sq | Df | F value | Pr(>F) |
|---|---|---|---|---|
| (Intercept) | 57189.5779 | 1 | 183.2956 | 0.0000 |
| sesev_fv_t1 | 263557.1698 | 1 | 844.7145 | 0.0000 |
| Framing | 1222.8893 | 1 | 3.9194 | 0.0480 |
| cont1 | 40.0779 | 1 | 0.1285 | 0.7201 |
| cont2 | 0.5846 | 1 | 0.0019 | 0.9655 |
| sesev_fv_t1:Framing | 1411.1856 | 1 | 4.5229 | 0.0336 |
| sesev_fv_t1:cont1 | 609.9385 | 1 | 1.9549 | 0.1623 |
| Framing:cont1 | 53.6660 | 1 | 0.1720 | 0.6784 |
| sesev_fv_t1:cont2 | 51.2097 | 1 | 0.1641 | 0.6855 |
| Framing:cont2 | 100.9352 | 1 | 0.3235 | 0.5696 |
| sesev_fv_t1:Framing:cont1 | 156.2693 | 1 | 0.5009 | 0.4793 |
| sesev_fv_t1:Framing:cont2 | 152.7560 | 1 | 0.4896 | 0.4842 |
| Residuals | 377528.9467 | 1210 | NA | NA |

| | eta.sq | eta.sq.part |
|---|---|---|
| sesev_fv_t1 | 0.3935 | 0.4111 |
| Framing | 0.0018 | 0.0032 |
| cont1 | 0.0001 | 0.0001 |
| cont2 | 0.0000 | 0.0000 |
| sesev_fv_t1:Framing | 0.0021 | 0.0037 |
| sesev_fv_t1:cont1 | 0.0009 | 0.0016 |
| Framing:cont1 | 0.0001 | 0.0001 |
| sesev_fv_t1:cont2 | 0.0001 | 0.0001 |
| Framing:cont2 | 0.0002 | 0.0003 |
| sesev_fv_t1:Framing:cont1 | 0.0002 | 0.0004 |
| sesev_fv_t1:Framing:cont2 | 0.0002 | 0.0004 |

#### Supplementary Materials 1.2: Perceived Booster Risk

##### Summary

Perception of risk to participant's health associated with receiving the vaccines was recorded pre- and post-manipulation. The main effect of Framing did not reach statistical significance ($F(1, 1210)=1.04$, $p=.308$, *np2*=.001). There was a Familiarity by Baseline Risk Perception interaction ($F(2, 1210)=4.82$, $p=.008$, *np2*=.008), evident in both contrasts (Baseline Risk by Contrast1: $F(1, 1210)=4.28$, $p=.039$, *np2*=.004 | Baseline Risk by Contrast2: $F(1, 1210)=6.03$, $p=.014$, *np2*=.005). No other statistically significant main effects or interactions were observed (all *ps*>.10). As depicted in Supplementary Figure 1b, there was limited difference between Familiarity conditions when the perception of risk was low at baseline. At high levels of Baseline Risk, Post-Manipulation Risk Perception was lowest for the previously experienced vaccine and highest for the Unfamiliar vaccine.

##### Perceived Booster Risk: Statistical Models

- serisk_fv_t2 (perceived risk associated with the framed vaccine post-manipulation)
- sesev_fv_t1 (perceived risk associated with the framed vaccine at baseline)

##### Main Effects Model

| | Sum Sq | Df | F value | Pr(>F) |
|---|---|---|---|---|
| (Intercept) | 81175.1304 | 1 | 193.8057 | 0.0000 |
| serisk_fv_t1 | 215659.8866 | 1 | 514.8883 | 0.0000 |
| Framing | 436.2962 | 1 | 1.0417 | 0.3076 |
| Familiarity | 894.8706 | 2 | 1.0683 | 0.3439 |
| serisk_fv_t1:Framing | 21.8767 | 1 | 0.0522 | 0.8193 |
| serisk_fv_t1:Familiarity | 4039.4547 | 2 | 4.8221 | 0.0082 |
| Framing:Familiarity | 74.8987 | 2 | 0.0894 | 0.9145 |
| serisk_fv_t1:Framing:Familiarity | 919.5879 | 2 | 1.0978 | 0.3340 |
| Residuals | 506805.9582 | 1210 | NA | NA |

| | eta.sq | eta.sq.part |
|---|---|---|
| serisk_fv_t1 | 0.2885 | 0.2985 |
| Framing | 0.0006 | 0.0009 |
| Familiarity | 0.0012 | 0.0018 |
| serisk_fv_t1:Framing | 0.0000 | 0.0000 |
| serisk_fv_t1:Familiarity | 0.0054 | 0.0079 |
| Framing:Familiarity | 0.0001 | 0.0001 |
| serisk_fv_t1:Framing:Familiarity | 0.0012 | 0.0018 |

**Model with pre-registered contrasts for Vaccine Familiarity**

| | Sum Sq | Df | F value | Pr(>F) |
|---|---|---|---|---|
| (Intercept) | 81175.1304 | 1 | 193.8057 | 0.0000 |
| serisk_fv_t1 | 215659.8866 | 1 | 514.8883 | 0.0000 |
| Framing | 436.2962 | 1 | 1.0417 | 0.3076 |
| cont1 | 109.4773 | 1 | 0.2614 | 0.6093 |
| cont2 | 754.8287 | 1 | 1.8022 | 0.1797 |
| serisk_fv_t1:Framing | 21.8767 | 1 | 0.0522 | 0.8193 |
| serisk_fv_t1:cont1 | 1791.4699 | 1 | 4.2771 | 0.0388 |
| Framing:cont1 | 11.7394 | 1 | 0.0280 | 0.8671 |
| serisk_fv_t1:cont2 | 2526.0142 | 1 | 6.0309 | 0.0142 |
| Framing:cont2 | 65.7193 | 1 | 0.1569 | 0.6921 |
| serisk_fv_t1:Framing:cont1 | 708.3207 | 1 | 1.6911 | 0.1937 |
| serisk_fv_t1:Framing:cont2 | 267.7285 | 1 | 0.6392 | 0.4242 |
| Residuals | 506805.9582 | 1210 | NA | NA |

| | eta.sq | eta.sq.part |
|---|---|---|
| serisk_fv_t1 | 0.2885 | 0.2985 |
| Framing | 0.0006 | 0.0009 |
| cont1 | 0.0001 | 0.0002 |
| cont2 | 0.0010 | 0.0015 |
| serisk_fv_t1:Framing | 0.0000 | 0.0000 |
| serisk_fv_t1:cont1 | 0.0024 | 0.0035 |
| Framing:cont1 | 0.0000 | 0.0000 |
| serisk_fv_t1:cont2 | 0.0034 | 0.0050 |
| Framing:cont2 | 0.0001 | 0.0001 |
| serisk_fv_t1:Framing:cont1 | 0.0009 | 0.0014 |
| serisk_fv_t1:Framing:cont2 | 0.0004 | 0.0005 |

#### Supplementary Materials 1.3: Booster Acceptance (Satisfaction, Happiness, and Anxiety)

##### Summary

At pre- and post-manipulation, participants were asked to imagine that the framed vaccine was their only option for a booster and to rate their Satisfaction, Happiness and Anxiety regarding this outcome. In order to reduce the data for analysis, the three variables were entered into a principal components analysis. Component loading suggested that Satisfaction and Happiness loaded onto a similar latent construct, while Anxiety was associated with another. Therefore, Satisfaction and Happiness were averaged together for the subsequent analyses, while Anxiety was treated separately. Two ANCOVAs were run on these outcomes. The first revealed a main effect of Framing ($F(1, 1210)=5.66$, $p=.018$, *np2*=.005), a two-way Framing by Familiarity interaction ($F(2, 1210)=3.63$, $p=.027$, *np2*=.006) and a three-way Baseline Satisfaction/Happiness by Framing by Familiarity interaction ($F(2, 1210)=3.43$, $p=.033$, *np2*=.006). This three-way interaction was significant for Contrast1 (Same vs. Other: $F(1, 1210)=4.11$, $p=.043$, *np2*=.003), but not Contrast2 (Familiar vs. Unfamiliar: $F(1, 1210)=2.56$, $p=.110$, *np2*=.002). As shown in Supplementary Figure 1c, for the Same and Unfamiliar vaccines, lowered satisfaction and happiness were generally associated with Negative Framing, particularly at low levels of Baseline Satisfaction/Happiness. There appeared to be no difference for the Familiar vaccine.

With respect to anxiety, there was a significant Baseline Anxiety by Framing interaction ($F(1, 1210)=5.53$, $p=.019$, *np2*=.005), but no other main effects or interactions (all $ps>.05$). As shown in Supplementary Figure 1d, Positive Framing appeared to increase Post-Manipulation Anxiety at very high levels of Baseline Anxiety, but was limited to the Same Condition (i.e., among participants who had high Baseline Anxiety regarding continuation with their previous vaccine type), with this interaction nearing significance (Anxiety by Framing by Contrast1: $F(1, 1210)=3.65$, $p=.056$, *np2*=.003).

##### Correlations and PCA for Framed Vaccine Side Effect Perceptions

##### (Satisfaction, Happiness and Anxiety)

**Corrrelations between variables** Variables are: switchsat_fv_t2 (satisfaction with the framed vaccine post-manipulation) switchhap_fv_t2 (happiness with the framed vaccine post-manipulation) switchanx_rev_fv_t2 (anxiety with the framed vaccine post-manipulation - variable reversed (+ve association))

```
##
##  Pearson's product-moment correlation
##
## data:  sat_dat$switchsat_fv_t2 and sat_dat$switchhap_fv_t2
## t = 78.489, df = 1220, p-value < 2.2e-16
## alternative hypothesis: true correlation is not equal to 0
## 95 percent confidence interval:
##  0.9038498 0.9224377
## sample estimates:
##       cor
## 0.9136199
```

```
##
##  Pearson's product-moment correlation
##
## data:  sat_dat$switchsat_fv_t2 and sat_dat$switchanx_rev_fv_t2
## t = 16.839, df = 1220, p-value < 2.2e-16
## alternative hypothesis: true correlation is not equal to 0
## 95 percent confidence interval:
##  0.3876229 0.4786818
## sample estimates:
##       cor
## 0.4342611
```

```
##
##  Pearson's product-moment correlation
##
## data:  sat_dat$switchhap_fv_t2 and sat_dat$switchanx_rev_fv_t2
## t = 17.55, df = 1220, p-value < 2.2e-16
## alternative hypothesis: true correlation is not equal to 0
## 95 percent confidence interval:
##  0.4030299 0.4926355
## sample estimates:
##       cor
## 0.4489607
```

##### PCA component loadings for post-manipulation measures

| | PC1 | PC2 | PC3 |
|---|---|---|---|
| switchsat_fv_t2 | 0.6135009 | -0.3383353 | -0.7135446 |
| switchhap_fv_t2 | 0.6338972 | -0.3278683 | 0.7004832 |
| switchanx_rev_fv_t2 | 0.4709469 | 0.8820610 | -0.0133223 |

##### PCA component loadings for pre-manipulation measures

| | PC1 | PC2 | PC3 |
|---|---|---|---|
| switchsat_fv_t1 | 0.6469202 | -0.2877156 | 0.7061969 |
| switchhap_fv_t1 | 0.6539002 | -0.2671390 | -0.7078498 |
| switchanx_rev_fv_t1 | 0.3923122 | 0.9197045 | 0.0153197 |

**Satisfaction and Happiness combined: Statistical Models**

- av_sathap_t2 (average satisfaction and happiness associated with the framed vaccine post-manipulation)
- av_sathap_t1 (average satisfaction and happiness associated with the framed vaccine at baseline)

**Main Effects Model**

| | Sum Sq | Df | F value | Pr(>F) |
|---|---|---|---|---|
| (Intercept) | 68464.8110 | 1 | 177.3429 | 0.0000 |
| av_sathap_t1 | 629741.7591 | 1 | 1631.2060 | 0.0000 |
| Framing | 2184.7699 | 1 | 5.6592 | 0.0175 |
| Familiarity | 1042.8219 | 2 | 1.3506 | 0.2595 |
| av_sathap_t1:Framing | 849.7667 | 1 | 2.2011 | 0.1382 |
| av_sathap_t1:Familiarity | 1420.4558 | 2 | 1.8397 | 0.1593 |
| Framing:Familiarity | 2803.5778 | 2 | 3.6310 | 0.0268 |
| av_sathap_t1:Framing:Familiarity | 2651.9911 | 2 | 3.4347 | 0.0325 |
| Residuals | 467131.3843 | 1210 | NA | NA |

| | eta.sq | eta.sq.part |
|---|---|---|
| av_sathap_t1 | 0.5173 | 0.5741 |
| Framing | 0.0018 | 0.0047 |
| Familiarity | 0.0009 | 0.0022 |
| av_sathap_t1:Framing | 0.0007 | 0.0018 |
| av_sathap_t1:Familiarity | 0.0012 | 0.0030 |
| Framing:Familiarity | 0.0023 | 0.0060 |
| av_sathap_t1:Framing:Familiarity | 0.0022 | 0.0056 |

**Model with pre-registered contrasts for Vaccine Familiarity**

| | Sum Sq | Df | F value | Pr(>F) |
|---|---|---|---|---|
| (Intercept) | 68464.8110 | 1 | 177.3429 | 0.0000 |
| av_sathap_t1 | 629741.7591 | 1 | 1631.2060 | 0.0000 |
| Framing | 2184.7699 | 1 | 5.6592 | 0.0175 |
| cont1 | 89.8365 | 1 | 0.2327 | 0.6296 |
| cont2 | 949.0046 | 1 | 2.4582 | 0.1172 |
| av_sathap_t1:Framing | 849.7667 | 1 | 2.2011 | 0.1382 |
| av_sathap_t1:cont1 | 7.8678 | 1 | 0.0204 | 0.8865 |
| Framing:cont1 | 1323.4860 | 1 | 3.4282 | 0.0643 |
| av_sathap_t1:cont2 | 1405.2058 | 1 | 3.6399 | 0.0566 |
| Framing:cont2 | 1461.2307 | 1 | 3.7850 | 0.0519 |
| av_sathap_t1:Framing:cont1 | 1588.5089 | 1 | 4.1147 | 0.0427 |
| av_sathap_t1:Framing:cont2 | 987.7901 | 1 | 2.5587 | 0.1100 |
| Residuals | 467131.3843 | 1210 | NA | NA |

| | eta.sq | eta.sq.part |
|---|---|---|
| av_sathap_t1 | 0.5173 | 0.5741 |
| Framing | 0.0018 | 0.0047 |
| cont1 | 0.0001 | 0.0002 |
| cont2 | 0.0008 | 0.0020 |
| av_sathap_t1:Framing | 0.0007 | 0.0018 |
| av_sathap_t1:cont1 | 0.0000 | 0.0000 |
| Framing:cont1 | 0.0011 | 0.0028 |
| av_sathap_t1:cont2 | 0.0012 | 0.0030 |
| Framing:cont2 | 0.0012 | 0.0031 |
| av_sathap_t1:Framing:cont1 | 0.0013 | 0.0034 |
| av_sathap_t1:Framing:cont2 | 0.0008 | 0.0021 |

**Anxiety: Statistical Models**

**Main Effects Model**

| | Sum Sq | Df | F value | Pr(>F) |
|---|---|---|---|---|
| (Intercept) | 80477.3132 | 1 | 140.8544 | 0.0000 |
| switchanx_fv_t1 | 584744.8174 | 1 | 1023.4421 | 0.0000 |
| Framing | 1449.0382 | 1 | 2.5362 | 0.1115 |
| Familiarity | 1743.2143 | 2 | 1.5255 | 0.2179 |
| switchanx_fv_t1:Framing | 3160.2594 | 1 | 5.5312 | 0.0188 |
| switchanx_fv_t1:Familiarity | 524.9404 | 2 | 0.4594 | 0.6318 |
| Framing:Familiarity | 318.5277 | 2 | 0.2787 | 0.7568 |
| switchanx_fv_t1:Framing:Familiarity | 2257.7873 | 2 | 1.9758 | 0.1391 |
| Residuals | 691334.8598 | 1210 | NA | NA |

| | eta.sq | eta.sq.part |
|---|---|---|
| switchanx_fv_t1 | 0.4358 | 0.4582 |
| Framing | 0.0011 | 0.0021 |
| Familiarity | 0.0013 | 0.0025 |
| switchanx_fv_t1:Framing | 0.0024 | 0.0046 |
| switchanx_fv_t1:Familiarity | 0.0004 | 0.0008 |
| Framing:Familiarity | 0.0002 | 0.0005 |
| switchanx_fv_t1:Framing:Familiarity | 0.0017 | 0.0033 |

**Model with pre-registered contrasts for Vaccine Familiarity**

| | Sum Sq | Df | F value | Pr(>F) |
|---|---|---|---|---|
| (Intercept) | 80477.3132 | 1 | 140.8544 | 0.0000 |
| switchanx_fv_t1 | 584744.8174 | 1 | 1023.4421 | 0.0000 |
| Framing | 1449.0382 | 1 | 2.5362 | 0.1115 |
| cont1 | 1041.1486 | 1 | 1.8223 | 0.1773 |
| cont2 | 788.8024 | 1 | 1.3806 | 0.2402 |
| switchanx_fv_t1:Framing | 3160.2594 | 1 | 5.5312 | 0.0188 |
| switchanx_fv_t1:cont1 | 4.6664 | 1 | 0.0082 | 0.9280 |
| Framing:cont1 | 174.0626 | 1 | 0.3047 | 0.5811 |
| switchanx_fv_t1:cont2 | 518.5455 | 1 | 0.9076 | 0.3409 |
| Framing:cont2 | 160.4686 | 1 | 0.2809 | 0.5962 |
| switchanx_fv_t1:Framing:cont1 | 2086.2670 | 1 | 3.6515 | 0.0563 |
| switchanx_fv_t1:Framing:cont2 | 152.6742 | 1 | 0.2672 | 0.6053 |
| Residuals | 691334.8598 | 1210 | NA | NA |

| | eta.sq | eta.sq.part |
|---|---|---|
| switchanx_fv_t1 | 0.4358 | 0.4582 |
| Framing | 0.0011 | 0.0021 |
| cont1 | 0.0008 | 0.0015 |
| cont2 | 0.0006 | 0.0011 |
| switchanx_fv_t1:Framing | 0.0024 | 0.0046 |
| switchanx_fv_t1:cont1 | 0.0000 | 0.0000 |
| Framing:cont1 | 0.0001 | 0.0003 |
| switchanx_fv_t1:cont2 | 0.0004 | 0.0007 |
| Framing:cont2 | 0.0001 | 0.0002 |
| switchanx_fv_t1:Framing:cont1 | 0.0016 | 0.0030 |
| switchanx_fv_t1:Framing:cont2 | 0.0001 | 0.0002 |

#### Supplementary Materials 1.4.1: Judgements Regarding Side Effect Prevalence

##### Summary

Participants were required to classify 14 side effects from their assigned PIL (see tables in 1.4.2 below) into verbal prevalence categories, as well as provide a frequency estimate regarding how many people, in 100,000, they thought would experience each side effect if they received the framed vaccine. Eleven side effects were associated with discrete prevalence categories. Three were presented in the PILs as of 'unknown prevalence'. Raw frequency estimates of the latter were analysed separately to test whether framing alters perceptions regarding prevalence when no information is provided. For side effects with a concrete category, the percentage of side effects correctly assigned across verbal prevalence categories, and frequency bands, were computed as outcome measures. Frequency estimates were judged correct based on the following frequency bands (see Berry, Raynor & Knapp, 2003): Very Common = 10,001–100,000 (>10%); Common = 1001–10,000 (>1–10%); Uncommon = 101–1000 (>0.1–1%); Rare = 11–100 (>0.01–0.1%); Very Rare = 0–10 (0–0.01%). ANOVAs were performed with Framing and Familiarity as factors.

##### Results

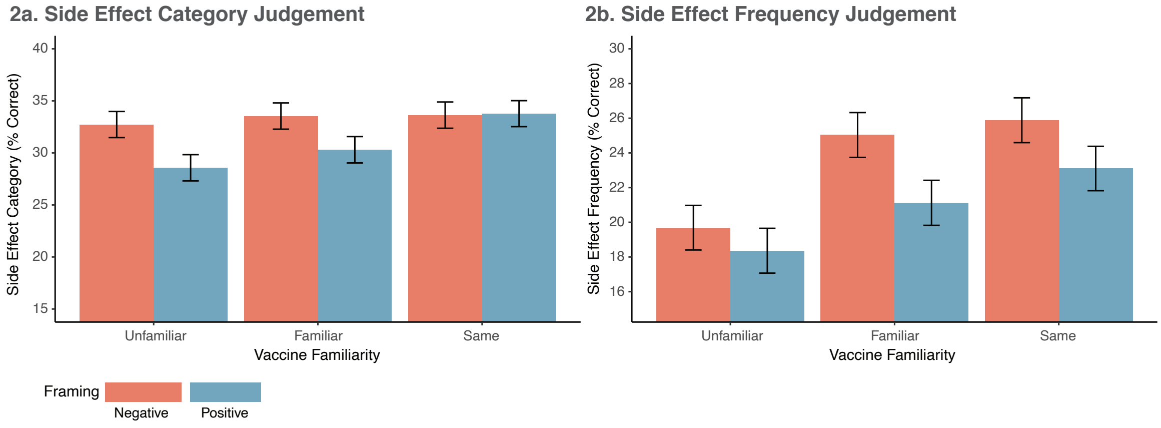


Supplementary Figure 2: Mean pearcentage of correct category (2a) and frequency (2b) judgements. All error bars represent +/- 1 SEM.

##### Verbal Prevalence Categories

As depicted in Supplementary Figure 2a, a main effect of Framing ($F(1, 1216)$=5.54, $p$=.019, *np2*=.005) was observed, while Familiarity neared significance ($F(2, 1216)$=2.98, $p$=.051, *np2*=.005). This was driven by Contrast1 only (Same vs. Other: $F(1, 1216)$=4.93, $p$=.027, *np2*=.004). While accuracy was generally low, fewer accurate classifications were made following Positive Framing ($M$=30.9%, $SEM$=0.73, 95% CIs [29.46, 32.31]), relative to Negative Framing ($M$=33.3%, $SEM$=0.73, 95% CIs [31.87, 43.73]). While the reduction in accuracy associated with positive, relative to negative, framing appeared to decrease with familiarity, the Framing by Contrast1(Same vs. Other) interaction did not reach statistical significance ($F(1, 1216)$=3.11, $p$=.078, *np2*=.003).

**Verbal Prevalence Categories: Statistical Models**

Overall_CategoryPercent = percentage of correctly identified categories

**Main Effects Model**

| | Sum Sq | Df | F value | Pr(>F) |
|---|---|---|---|---|
| (Intercept) | 1258379.599 | 1 | 3900.3990 | 0.0000 |
| Framing | 1786.274 | 1 | 5.5366 | 0.0188 |
| Familiarity | 1923.618 | 2 | 2.9812 | 0.0511 |
| Framing:Familiarity | 1047.111 | 2 | 1.6228 | 0.1978 |
| Residuals | 392316.166 | 1216 | NA | NA |

| | eta.sq | eta.sq.part |
|---|---|---|
| Framing | 0.0045 | 0.0045 |
| Familiarity | 0.0048 | 0.0049 |
| Framing:Familiarity | 0.0026 | 0.0027 |

**Model with pre-registered contrasts for Vaccine Familiarity**

| | Sum Sq | Df | F value | Pr(>F) |
|---|---|---|---|---|
| (Intercept) | 1258379.5992 | 1 | 3900.3990 | 0.0000 |
| Framing | 1786.2735 | 1 | 5.5366 | 0.0188 |
| cont1 | 1590.3548 | 1 | 4.9294 | 0.0266 |
| cont2 | 329.1191 | 1 | 1.0201 | 0.3127 |
| Framing:cont1 | 1003.2909 | 1 | 3.1097 | 0.0781 |
| Framing:cont2 | 42.6316 | 1 | 0.1321 | 0.7163 |
| Residuals | 392316.1662 | 1216 | NA | NA |

| | eta.sq | eta.sq.part |
|---|---|---|
| Framing | 0.0045 | 0.0045 |
| cont1 | 0.0040 | 0.0040 |
| cont2 | 0.0008 | 0.0008 |
| Framing:cont1 | 0.0025 | 0.0026 |
| Framing:cont2 | 0.0001 | 0.0001 |

##### Frequency Estimates

A main effect of Framing ($F(1, 1216)=6.45$, $p=.011$, $np2=.005$) and Familiarity ($F(2, 1216)=9.71$, $p=.0001$, $np2=.016$) was observed. As depicted in Supplementary Figure 2b, accuracy was reduced with Positive Framing, but increased with Familiarity. Contrast1 (Same vs. Other: $F(1, 1216)=9.52$, $p=.002$, $np2=.008$) and Contrast2 (Familiar vs. Unfamiliar: $F(1, 1216)=9.84$, $p=.002$, $np2=.008$) reached significance. Overall, participants frequency estimates were less accurate following positive framing ($M=20.7\%$, $SEM=0.74$, 95% CIs [19.40, 22.32]) relative to negative framing ($M=23.54\%$, $SEM=0.74$, 95% CIs [22.08, 25.00]). Framing did not interact with the main effect of Familiarity, or the orthogonal contrasts (all $ps>.30$).

Neither Framing nor Familiarity had any effect on frequency estimates of side effects of 'unknown' frequency (all $ps>.16$). Those in the Positive Framing conditions estimated that these side effects would be experienced by 1,848/100,000 people on average, and those in the Negative Frame as 2,004/100,000.

##### Frequency Estimates: Statistical Models

Overall_FrequencyPercent = percentage of correctly estimated frequency brackets

##### Main Effects Model

| | Sum Sq | Df | F value | Pr(>F) |
|---|---|---|---|---|
| (Intercept) | 602115.9981 | 1 | 1777.6846 | 0.0000 |
| Framing | 2185.5526 | 1 | 6.4526 | 0.0112 |
| Familiarity | 6577.8779 | 2 | 9.7102 | 0.0001 |
| Framing:Familiarity | 340.6265 | 2 | 0.5028 | 0.6049 |
| Residuals | 411868.9225 | 1216 | NA | NA |

| | eta.sq | eta.sq.part |
|---|---|---|
| Framing | 0.0052 | 0.0053 |
| Familiarity | 0.0156 | 0.0157 |
| Framing:Familiarity | 0.0008 | 0.0008 |

##### Model with pre-registered contrasts for Vaccine Familiarity

| | Sum Sq | Df | F value | Pr(>F) |
|---|---|---|---|---|
| (Intercept) | 602115.9981 | 1 | 1777.6846 | 0.0000 |
| Framing | 2185.5526 | 1 | 6.4526 | 0.0112 |
| cont1 | 3225.6804 | 1 | 9.5235 | 0.0021 |
| cont2 | 3333.4302 | 1 | 9.8416 | 0.0017 |
| Framing:cont1 | 1.8231 | 1 | 0.0054 | 0.9415 |
| Framing:cont2 | 338.6588 | 1 | 0.9999 | 0.3175 |
| Residuals | 411868.9225 | 1216 | NA | NA |

| | eta.sq | eta.sq.part |
|---|---|---|
| Framing | 0.0052 | 0.0053 |
| cont1 | 0.0077 | 0.0078 |
| cont2 | 0.0079 | 0.0080 |
| Framing:cont1 | 0.0000 | 0.0000 |
| Framing:cont2 | 0.0008 | 0.0008 |

**Frequency estimates for side effect of 'unknown prevalence'**

**Main Effects Model**

| | Sum Sq | Df | F value | Pr(>F) |
|---|---|---|---|---|
| (Intercept) | 4533390495 | 1 | 29.5304 | 0.0000 |
| Framing | 7457785 | 1 | 0.0486 | 0.8256 |
| Familiarity | 537592293 | 2 | 1.7509 | 0.1740 |
| Framing:Familiarity | 119664905 | 2 | 0.3897 | 0.6773 |
| Residuals | 186675244185 | 1216 | NA | NA |

| | eta.sq | eta.sq.part |
|---|---|---|
| Framing | 0.0000 | 0.0000 |
| Familiarity | 0.0029 | 0.0029 |
| Framing:Familiarity | 0.0006 | 0.0006 |

#### Supplementary Materials 1.4.2: Side effects presented from each PIL

The following presents the 14 side effects associated with each vaccine type (i.e., AstraZeneca, Pfizer, Moderna).

| AstraZeneca Side Effects | |
|---|---|
| ***Category*** | ***Side Effect ($N = 11$)*** |
| Very Common | Injection site pain |
| Very Common | Tiredness (fatigue) |
| Very Common | Muscle ache/pain |
| Very Common | Chills |
| Common | Injection site redness |
| Common | Vomiting |
| Uncommon | Diarrhea |
| Uncommon | Enlarged lymph nodes (swelling of underarm glands) |
| Uncommon | Dizziness |
| Very Rare | Blood clots often in unusual locations (e.g. brain, bowel, liver, spleen) in combination with low level of blood platelets |
| Very Rare | Serious nerve inflammation, which may cause paralysis and difficulty breathing (Guillain-Barré syndrome) |

| AstraZeneca Unknown Side Effects | |
|---|---|
| ***Category*** | ***Side Effect ($N = 3$)*** |
| Not known | Severe allergic reaction (anaphylaxis) |
| Not known | hypersensitivity |
| Not known | capillary leak syndrome (a condition causing fluid leakage from small blood vessels) |

| Pfizer Side Effects | |
|---|---|
| **Category** | **Side Effect (N = 11)** |
| Very Common | Injection site pain |
| Very Common | Tiredness (fatigue) |
| Very Common | Muscle ache/pain |
| Very Common | Chills |
| Common | Injection site redness |
| Common | Vomiting |
| Uncommon | Diarrhea |
| Uncommon | Enlarged lymph nodes (swelling of underarm glands) |
| Uncommon | Insomnia |
| Rare | Temporary one-sided facial drooping |
| Rare | Swelling of the face |

| Pfizer Unknown Side Effects | |
|---|---|
| **Category** | **Side Effect (N = 3)** |
| Not known | Severe allergic reaction (anaphylaxis) |
| Not known | Inflammation of the heart (myocarditis or pericarditis) |
| Not known | Extensive swelling of the vaccinated limb |

| Moderna Side Effects | |
|---|---|
| **Category** | **Side Effect (N = 11)** |
| Very Common | Injection site pain |
| Very Common | Tiredness (fatigue) |
| Very Common | Muscle ache/pain |
| Very Common | Chills |
| Common | Injection site redness |
| Common | Rash |
| Common | Injection site hives |
| Uncommon | injection site itching |
| Rare | Temporary one-sided facial drooping |
| Rare | Swelling of the face |
| Rare | Decreased sense of touch or sensation |

| Moderna Unknown Side Effects | |
|---|---|
| **Category** | **Side Effect (N = 3)** |
| Not known | Severe allergic reaction (anaphylaxis) |
| Not known | Hypersensitivity |
| Not known | Inflammation of the heart (myocarditis or pericarditis) |

### Supplementary Materials 2

#### Pre-analysis data cleaning

**Participants (N = 237) were excluded based on meeting one or more of the following pre-registered quality control checks:**

1. Completed the study quicker than would be expected given a reasonable reading rate (8 minutes, 5th percentile of soft-launch data)
2. Failed to answer attention questions appropriately (e.g., place slider in X position)
3. Provided discrepant data during screening and the main survey regarding the COVID-19 vaccine they had previously received (i.e., AstraZeneca or Pfizer)
4. Identified as having experienced side effects to previous COVID-19 vaccines, then did not select any side effects
5. Spent more than 5 minutes on the 2-minute timed manipulation page without proceeding
6. Identified as having previously received a specific COVID-19 vaccine, then stated they had never heard of that vaccine

### Supplementary Materials 3

#### Wording used for the Positive and Negative Attribute Frame

| Negative Attribute Frame (Standard EU Wording) | Positive Attribute Frame | Positive Attribute Frame (Accompanying Infographic) | |
|---|---|---|---|
| **Very common** (may affect more than 1 in 10 people) | **Very common** (9 in 10 or fewer people may ***not be affected***) | 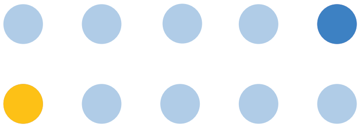 | **Very Common:** 9 in 10 or fewer **not affected** (light to dark blue circles) |
| **Common** (may affect up to 1 in 10 people) | **Common** (90 in 100 or more people may ***not be affected***) | 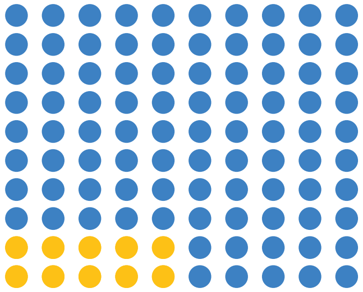 | **Common:** 90 in 100 (blue circles) or more **not affected** |
| **Uncommon** (may affect up to 1 in 100 people) | **Uncommon** (990 in 1,000 or more people may ***not be affected***) | 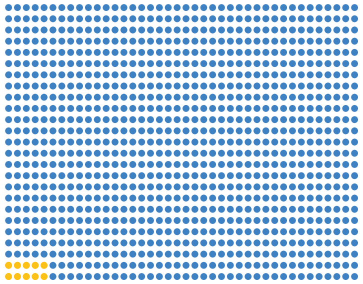 | **Uncommon:** 990 in 1,000 (blue circles) or more **not affected** |
| **Rare** (may affect up to 1 in 1,000 people) | **Rare** (9,990 in 10,000 or more people may ***not be affected***) | 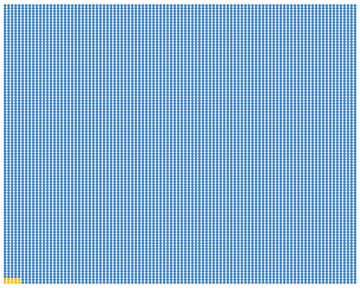 | **Rare:** 9,990 in 10,000 (blue circles) or more **not affected** |
| **Very Rare** (may affect up to 1 in 10,000 people) | **Very Rare** (99,990 in 100,000 or more people may ***not be affected***) | 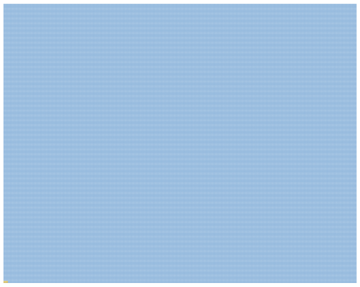 | **Very Rare:** 99,990 in 100,000 (blue circles) or more **not affected** |

### Supplementary Materials 4

#### Patient Information Leaflets (PILs) employed in the present experiment

The following pages contain the PILs used to present side effect information.

High fidelity versions of these documents were embedded in Qualtrics.

### Negative PIL (AstraZeneca Vaccine)

**Package leaflet: Information for the user**

**Vaxzevria suspension for injection**
COVID-19 Vaccine (ChAdOx1-S [recombinant])

▼This medicine is subject to additional monitoring. This will allow quick identification of new safety information. You can help by reporting any side effects you may get.

**Read all of this leaflet carefully before the vaccine is given because it contains important information for you.**

**What Vaxzevria is and what it is used for**

Vaxzevria is used for preventing COVID-19 caused by the SARS-CoV-2 virus.

Vaxzevria is given to adults aged 18 years and older.

The vaccine causes the immune system (the body's natural defences) to produce antibodies and specialised white blood cells that work against the virus, so giving protection against COVID-19.
None of the ingredients in this vaccine can cause COVID-19

**Possible side effects**

Like all medicines, this vaccine can cause side effects, although not everybody gets them.

**The following side effects may occur with Vaxzevria:**

**Very Common** (may affect more than 1 in 10 people)

- tenderness, pain, warmth, itching, or bruising where the injection is given
- feeling tired (fatigue) or generally feeling unwell
- chills or feeling feverish
- headache
- feeling sick (nausea)
- joint pain or muscle ache

**Common** (may affect up to 1 in 10 people)

- swelling or redness where the injection is given
- fever (≥38°C)
- being sick (vomiting) or diarrhoea
- low level of blood platelets
- pain in legs or arms
- flu-like symptoms, such as high temperature, sore throat, runny nose, cough and chills
- physical weakness or lack of energy

**Uncommon** (may affect up to 1 in 100 people)

- sleepiness or feeling dizzy
- abdominal pain or decreased appetite
- enlarged lymph nodes
- excessive sweating, itchy skin, rash or hives
- sleepiness or deep unresponsiveness and inactivity

**Very Rare** (may affect up to 1 in 10,000 people)

- blood clots often in unusual locations (e.g. brain, bowel, liver, spleen) in combination with low level of blood platelets
- serious nerve inflammation, which may cause paralysis and difficulty breathing (Guillain-Barré syndrome [GBS])

**Not known** (cannot be estimated from the available data)

- severe allergic reaction (anaphylaxis)
- hypersensitivity
- rapid swelling under the skin in areas such as the face, lips, mouth and throat (which may cause difficulty in swallowing or breathing)
- capillary leak syndrome (a condition causing fluid leakage from small blood vessels)

### Postive PIL (AstraZeneca Vaccine)

**Package leaflet: Information for the user**

**Vaxzevria suspension for injection**
COVID-19 Vaccine (ChAdOx1-S [recombinant])

▼This medicine is subject to additional monitoring. This will allow quick identification of new safety information. You can help by reporting any side effects you may get.

**Read all of this leaflet carefully before the vaccine is given because it contains important information for you.**

**What Vaxzevria is and what it is used for**

Vaxzevria is used for preventing COVID-19 caused by the SARS-CoV-2 virus.

Vaxzevria is given to adults aged 18 years and older.

The vaccine causes the immune system (the body's natural defences) to produce antibodies and specialised white blood cells that work against the virus, so giving protection against COVID-19.
None of the ingredients in this vaccine can cause COVID-19

**In the images below:**
Blue circles represent people **not affected**
Yellow circles represent people who **are affected**

**Possible side effects**

Like all medicines, this vaccine can cause side effects, although not everybody gets them.

**The following side effects may occur with Vaxzevria:**

**Very Common** (9 in 10 or fewer people may ***not be affected***)

- tenderness, pain, warmth, itching, or bruising where the injection is given
- feeling tired (fatigue) or generally feeling unwell
- chills or feeling feverish
- headache
- feeling sick (nausea)
- joint pain or muscle ache

**Very Common:** 9 in 10 or fewer **not affected** (light to dark blue circles)

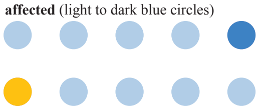

**Common** (90 in 100 or more people may ***not be affected***)

- swelling or redness where the injection is given
- fever (≥38°C)
- being sick (vomiting) or diarrhoea
- low level of blood platelets
- pain in legs or arms
- flu-like symptoms, such as high temperature, sore throat, runny nose, cough and chills
- physical weakness or lack of energy

**Common:** 90 in 100 (blue circles) or more **not affected**

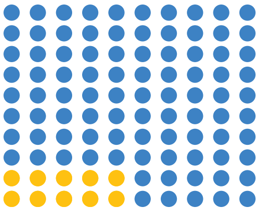

**Uncommon** (990 in 1,000 or more people may ***not be affected***)

- sleepiness or feeling dizzy
- abdominal pain or decreased appetite
- enlarged lymph nodes
- excessive sweating, itchy skin, rash or hives
- sleepiness or deep unresponsiveness and inactivity

**Uncommon:** 990 in 1,000 (blue circles) or more **not affected**

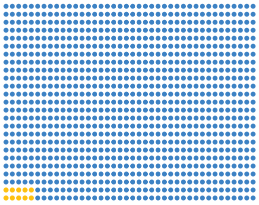

**Very Rare** (99,990 in 100,000 or more people may ***not be affected***)

- blood clots often in unusual locations (e.g. brain, bowel, liver, spleen) in combination with low level of blood platelets
- serious nerve inflammation, which may cause paralysis and difficulty breathing (Guillain-Barré syndrome [GBS])

**Very Rare:** 99,990 in 100,000 (blue circles) or more **not affected**

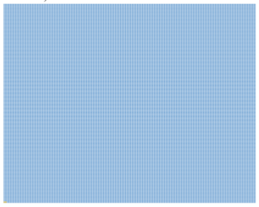

**Not known** (cannot be estimated from the available data)

- severe allergic reaction (anaphylaxis)
- hypersensitivity
- rapid swelling under the skin in areas such as the face, lips, mouth and throat (which may cause difficulty in swallowing or breathing)
- capillary leak syndrome (a condition causing fluid leakage from small blood vessels)

### Negative PIL (Pfizer Vaccine)

**Package leaflet: Information for the user**

**Comirnaty concentrate for dispersion for injection**
COVID-19 mRNA Vaccine (nucleoside modified)

▼This medicine is subject to additional monitoring. This will allow quick identification of new safety information. You can help by reporting any side effects you may get.

**Read all of this leaflet carefully before you receive this vaccine because it contains important information for you.**

**What Comirnaty is and what it is used for**

Comirnaty is a vaccine used for preventing COVID-19 caused by SARS-CoV-2 virus.

Comirnaty is given to adults and adolescents from 12 years of age and older.

The vaccine causes the immune system (the body's natural defences) to produce antibodies and blood cells that work against the virus, so giving protection against COVID-19.

As Comirnaty does not contain the virus to produce immunity, it cannot give you COVID-19.

**Possible side effects**

Like all vaccines, Comirnaty can cause side effects, although not everybody gets them.

**The following side effects may occur with Comirnaty:**

**Very common side effects** (may affect more than 1 in 10 people)

- injection site: pain, swelling
- tiredness
- headache
- muscle pain
- chills
- joint pain
- diarrhoea
- fever

Some of these side effects were slightly more frequent in adolescents 12 to 15 years than in adults.

**Common side effects** (may affect up to 1 in 10 people)

- injection site redness
- nausea
- vomiting

**Uncommon side effects** (may affect up to 1 in 100 people)

- enlarged lymph nodes
- feeling unwell
- arm pain
- insomnia
- injection site itching
- allergic reactions such as rash or itching

**Rare side effects** (may affect up to 1 in 1,000 people)

- temporary one sided facial drooping
- allergic reactions such as hives or swelling of the face

**Not known** (cannot be estimated from the available data)

- severe allergic reaction
- inflammation of the heart muscle (myocarditis) or inflammation of the lining outside the heart (pericarditis) which can result in breathlessness, palpitations or chest pain
- extensive swelling of the vaccinated limb
- swelling of the face (swelling of the face may occur in patients who have had facialdermatological fillers)

### Postive PIL (Pfizer Vaccine)

**Package leaflet: Information for the user**

**Comirnaty concentrate for dispersion for injection**
COVID-19 mRNA Vaccine (nucleoside modified)

▼This medicine is subject to additional monitoring. This will allow quick identification of new safety information. You can help by reporting any side effects you may get.

**Read all of this leaflet carefully before you receive this vaccine because it contains important information for you.**

**What Comirnaty is and what it is used for**

Comirnaty is a vaccine used for preventing COVID-19 caused by SARS-CoV-2 virus.

Comirnaty is given to adults and adolescents from 12 years of age and older.

The vaccine causes the immune system (the body's natural defences) to produce antibodies and blood cells that work against the virus, so giving protection against COVID-19.

As Comirnaty does not contain the virus to produce immunity, it cannot give you COVID-19.

**Possible side effects**

Like all vaccines, Comirnaty can cause side effects, although not everybody gets them.

**The following side effects may occur with Comirnaty:**

**Very common side effects** (9 in 10 or fewer people may ***not be affected***)

- injection site: pain, swelling
- tiredness
- headache
- muscle pain
- chills
- joint pain
- diarrhoea
- fever

**Very Common:** 9 in 10 or fewer **not affected** (light to dark blue circles)

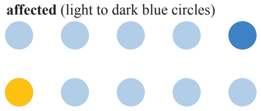

Some of these side effects were slightly more frequent in adolescents 12 to 15 years than in adults.

**Common side effects** (90 in 100 or more people may ***not be affected***)

- injection site redness
- nausea
- vomiting

**Common:** 90 in 100 (blue circles) or more **not affected**

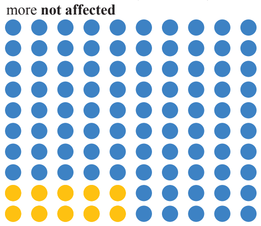

**Uncommon side effects** (990 in 1,000 or more people may ***not be affected***)

- enlarged lymph nodes
- feeling unwell
- arm pain
- insomnia
- injection site itching
- allergic reactions such as rash or itching

**Uncommon:** 990 in 1,000 (blue circles) or more **not affected**

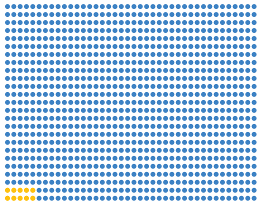

**Rare side effects** (9,990 in 10,000 or more people may ***not be affected***)

- temporary one sided facial drooping
- allergic reactions such as hives or swelling of the face

**Rare:** 9,990 in 10,000 (blue circles) or more **not affected**

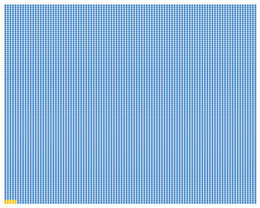

**Not known** (cannot be estimated from the available data)

- severe allergic reaction
- inflammation of the heart muscle (myocarditis) or inflammation of the lining outside the heart (pericarditis) which can result in breathlessness, palpitations or chest pain
- extensive swelling of the vaccinated limb
- swelling of the face (swelling of the face may occur in patients who have had facialdermatological fillers)

### Negative PIL (Moderna Vaccine)

**Package leaflet: Information for the user**

**Spikevax dispersion for injection**
COVID-19 mRNA Vaccine (nucleoside modified)

▼This medicine is subject to additional monitoring. This will allow quick identification of new safety information. You can help by reporting any side effects you may get.

**Read all of this leaflet carefully before you receive this vaccine because it contains important information for you.**

**What Spikevax is and what it is used for**

Spikevax is a vaccine used to prevent COVID-19 caused by SARS-CoV-2. It is given to adults and children aged 12 years and older. The active substance in Spikevax is mRNA encoding the SARS-CoV-2 Spike protein. The mRNA is embedded in SM-102 lipid nanoparticles.
As Spikevax does not contain the virus, it cannot give you COVID-19.

**How the vaccine works**
Spikevax stimulates the body's natural defences (immune system). The vaccine works by causing the body to produce protection (antibodies) against the virus that causes COVID-19.

**Possible side effects**

Like all medicines, this vaccine can cause side effects, although not everybody gets them.

**The following side effects may occur with Spikevax:**

**Very common** (may affect more than 1 in 10 people)

- swelling/tenderness of the underarm glands
- headache
- nausea
- vomiting
- muscle ache, joint aches, and stiffness
- pain or swelling at the injection site
- feeling very tired
- chills
- fever

**Common** (may affect up to 1 in 10 people)

- rash
- rash, redness, or hives at the injection site (some of which may occur some time after the
- injection)

**Uncommon** (may affect up to 1 in 100 people)

- itchiness at the injection site

**Rare** (may affect up to 1 in 1,000 people)

- temporary one-sided facial drooping (Bell's palsy)
- swelling of the face (Swelling of the face may occur in patients who have had facial cosmetic
- injections.)
- dizziness
- decreased sense of touch or sensation

**Frequency unknown** (cannot be estimated from the available data)

- severe allergic reactions with breathing difficulties (anaphylaxis)
- reaction of increased sensitivity or intolerance by the immune system (hypersensitivity)
- inflammation of the heart muscle (myocarditis) or inflammation of the lining outside the heart (pericarditis) which can result in breathlessness, palpitations or chest pain.

### Postive PIL (Moderna Vaccine)

**Package leaflet: Information for the user**

**Spikevax dispersion for injection**
COVID-19 mRNA Vaccine (nucleoside modified)

▼This medicine is subject to additional monitoring. This will allow quick identification of new safety information. You can help by reporting any side effects you may get.

**Read all of this leaflet carefully before you receive this vaccine because it contains important information for you.**

**What Spikevax is and what it is used for**

Spikevax is a vaccine used to prevent COVID-19 caused by SARS-CoV-2. It is given to adults and children aged 12 years and older. The active substance in Spikevax is mRNA encoding the SARS-CoV-2 Spike protein. The mRNA is embedded in SM-102 lipid nanoparticles.
As Spikevax does not contain the virus, it cannot give you COVID-19.

**How the vaccine works**
Spikevax stimulates the body's natural defences (immune system). The vaccine works by causing the body to produce protection (antibodies) against the virus that causes COVID-19.

**Possible side effects**

Like all medicines, this vaccine can cause side effects, although not everybody gets them.

**The following side effects may occur with Spikevax:**

**Very common** (9 in 10 or fewer people may ***not be affected***)

- swelling/tenderness of the underarm glands
- headache
- nausea
- vomiting
- muscle ache, joint aches, and stiffness
- pain or swelling at the injection site
- feeling very tired
- chills
- fever

**Very Common:** 9 in 10 or fewer **not affected** (light to dark blue circles)

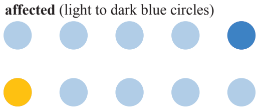

**Common** (90 in 100 or more people may ***not be affected***)

- rash
- rash, redness, or hives at the injection site (some of which may occur some time after the
- injection)

**Common:** 90 in 100 (blue circles) or more **not affected**

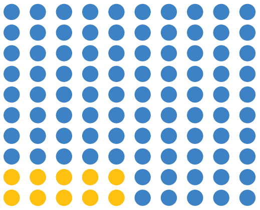

**Uncommon** (990 in 1,000 or more people may ***not be affected***)

- itchiness at the injection site

**Uncommon:** 990 in 1,000 (blue circles) or more **not affected**

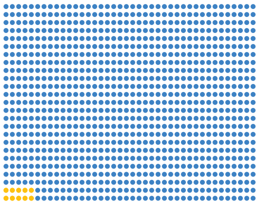

**Rare** (9,990 in 10,000 or more people may ***not be affected***)

- temporary one-sided facial drooping (Bell's palsy)
- swelling of the face (Swelling of the face may occur in patients who have had facial cosmetic
- injections.)
- dizziness
- decreased sense of touch or sensation

**Rare:** 9,990 in 10,000 (blue circles) or more **not affected**

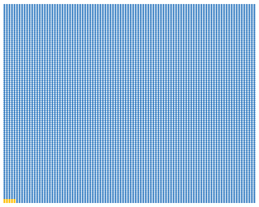

**Frequency unknown** (cannot be estimated from the available data)

- severe allergic reactions with breathing difficulties (anaphylaxis)
- reaction of increased sensitivity or intolerance by the immune system (hypersensitivity)
- inflammation of the heart muscle (myocarditis) or inflammation of the lining outside the heart (pericarditis) which can result in breathlessness, palpitations or chest pain.

### Supplementary Materials 5

#### Demographic Information (Full Sample)

##### Descriptive Statistics: Gender

Question wording:

"Which of the following most accurately describes you?"

Options:

- woman
- man
- non-binary/genderqueer/agender/gender fluid (non-binary)
- not sure
- prefer not to say
- other - let me type

| Gender | N |
|---|---|
| woman | 706 |
| man | 511 |
| non-binary | 3 |
| not sure | 1 |
| prefer not to say | 1 |

##### Descriptive Statistics: Education

Question wording:

"Please select your highest level of education:"

Options:

- GNVQ / GSVQ / GCSE/ SCE standard (GCSE)
- NVQ1, NVQ2 (NVQ1/2)
- NVQ3/ SCE Higher Grade/ Advanced GNVQ/ GCE A/AS or similar (A-Levels)
- NVQ4 / HNC / HND / Bachelor's degree or similar (Bachelors)
- NVQ5 or post-graduate diploma (Post-Graduate)
- Primary/Secondary school (no qualifications) (Primary/Secondary)
- Other (please specify)

| Education | N |
|---|---|
| Bachelors | 413 |
| A-Levels | 285 |
| GCSE | 255 |
| Post-Graduate | 121 |
| NVQ1/2 | 70 |
| Primary / Secondary | 53 |
| Other | 25 |

**Descriptive Statistics: Employment**

Question wording:

"Which of the following best describes you?"

Options:

- Employed full-time (full-time)
- Employed part-time (part-time)
- Full-time parenting / caring responsibilities (parent / carer)
- Retired
- Self employed (self-employed)
- Student
- Unemployed and not looking for a job / Long-term sick or disabled (unemployed not looking)
- Unemployed but looking for a job (unemployed looking)
- Other (not stated)

| Employment | N |
|---|---|
| full-time | 407 |
| retired | 321 |
| part-time | 153 |
| self-employed | 98 |
| unemployed (not looking) | 92 |
| parent / carer | 70 |
| unemployed (looking) | 38 |
| student | 24 |
| other | 19 |

**Descriptive Statistics: Ethnicity**

Question wording:

"Please select the option that best describes your ethnicity"

Options:

- Asian or Asian British - Bangladeshi
- Asian or Asian British - Indian (A/AB_I)
- Asian or Asian British - Pakistani (A/AB_P)
- Asian or Asian British - Any other Asian background (A/AB_Other)
- Black or Black British - African (B/BB_A)
- Black or Black British - Caribbean (B/BB_C)
- Black or Black British - Any other Black background
- Mixed - White and Black Caribbean (M/W&BC)
- Mixed - White and Black African
- Mixed - White and Asian (M/W&A)
- Mixed - Any other Mixed background
- Other ethnic groups - Chinese (OEG_C)
- Other ethnic groups - Any other ethnic group (ONS)
- White - British (W/B)
- White - Irish (W/I)
- White - Any other White background (W/O)

| Ethnicity | N |
| --- | --- |
| W/B | 1064 |
| W/O | 58 |
| A/AB_I | 19 |
| W/I | 16 |
| B/BB_A | 11 |
| M/W&BC | 10 |
| A/AB_P | 9 |
| OEG_C | 7 |
| A/AB_Other | 5 |
| B/BB_C | 5 |
| M/Other | 5 |
| A/AB_B | 4 |
| M/W&A | 4 |
| OEG | 3 |
| M/W&BA | 1 |
| ONS | 1 |

### Supplementary Materials 6

#### Demographic Information (by Experimental Condition)

##### Gender*Condition

| | man | non-binary | not sure | prefer not to say | woman |
|---|---|---|---|---|---|
| -ve familiar | 79 | 1 | 0 | 0 | 123 |
| -ve same | 81 | 0 | 0 | 0 | 122 |
| -ve unfamiliar | 87 | 1 | 0 | 1 | 116 |
| +ve familiar | 93 | 1 | 1 | 0 | 106 |
| +ve same | 90 | 0 | 0 | 0 | 117 |
| +ve unfamiliar | 81 | 0 | 0 | 0 | 122 |

##### Education*Condition

| | GCSE | NVQ1/2 | A-Levels | Bachelors | Post-Graduate | Other | Primary / Secondary |
|---|---|---|---|---|---|---|---|
| -ve familiar | 48 | 13 | 46 | 61 | 21 | 7 | 7 |
| -ve same | 44 | 10 | 50 | 66 | 14 | 4 | 15 |
| -ve unfamiliar | 44 | 11 | 47 | 71 | 27 | 2 | 3 |
| +ve familiar | 39 | 10 | 44 | 76 | 17 | 5 | 10 |
| +ve same | 36 | 16 | 53 | 65 | 21 | 5 | 11 |
| +ve unfamiliar | 44 | 10 | 45 | 74 | 21 | 2 | 7 |

##### Employment*Condition

| | full-time | part-time | parent / carer | other | retired | self-employed | student | unemployed (not looking) | unemployed (looking) |
|---|---|---|---|---|---|---|---|---|---|
| -ve familiar | 61 | 36 | 10 | 1 | 47 | 17 | 6 | 16 | 9 |
| -ve same | 63 | 28 | 13 | 2 | 52 | 19 | 4 | 16 | 6 |
| -ve unfamiliar | 73 | 29 | 10 | 2 | 46 | 21 | 5 | 15 | 4 |
| +ve familiar | 70 | 21 | 10 | 7 | 58 | 14 | 2 | 14 | 5 |
| +ve same | 68 | 16 | 10 | 3 | 66 | 16 | 3 | 17 | 8 |
| +ve unfamiliar | 72 | 23 | 17 | 4 | 52 | 11 | 4 | 14 | 6 |

**Ethnicity*Condition**

**Categories are:**

- 1 = "A/AB_Other"
- 2 = "A/AB_B"
- 3 = "A/AB_I"
- 4 = "A/AB_P"
- 5 = "B/BB_A"
- 6 = "B/BB_C"
- 7 = "M/Other"
- 8 = "M/W&A"
- 9 = "M/W&BA"
- 10 = "M/W&BC"
- 11 = "ONS"
- 12 = "OEG"
- 13 = "OEG_C"
- 14 = "W/O"
- 15 = "W/B"
- 16 = "W/I"

| | 1 | 2 | 3 | 4 | 5 | 6 | 7 | 8 | 9 | 10 | 11 | 12 | 13 | 14 | 15 | 16 |
|---|---|---|---|---|---|---|---|---|---|---|---|---|---|---|---|---|
| -ve familiar | 0 | 1 | 1 | 1 | 1 | 0 | 1 | 0 | 1 | 2 | 0 | 0 | 0 | 8 | 183 | 4 |
| -ve same | 2 | 2 | 3 | 1 | 3 | 1 | 0 | 2 | 0 | 0 | 0 | 0 | 2 | 10 | 174 | 3 |
| -ve unfamiliar | 1 | 0 | 4 | 3 | 2 | 0 | 2 | 0 | 0 | 2 | 0 | 0 | 3 | 14 | 170 | 4 |
| +ve familiar | 0 | 1 | 3 | 1 | 1 | 2 | 1 | 0 | 0 | 4 | 0 | 0 | 1 | 9 | 177 | 1 |
| +ve same | 0 | 0 | 5 | 0 | 3 | 2 | 1 | 2 | 0 | 1 | 1 | 2 | 0 | 8 | 181 | 1 |
| +ve unfamiliar | 2 | 0 | 3 | 3 | 1 | 0 | 0 | 0 | 0 | 1 | 0 | 1 | 1 | 9 | 179 | 3 |

**Months Since Last COVID Vaccine*Condition**

| | 0 | 1 | 2 | 3 | 4 | 5 | 6 | 7 | 8 | 9 | 10 | 11 | 13 |
|---|---|---|---|---|---|---|---|---|---|---|---|---|---|
| -ve familiar | 0 | 4 | 13 | 31 | 34 | 68 | 41 | 7 | 4 | 0 | 0 | 0 | 1 |
| -ve same | 0 | 6 | 14 | 24 | 30 | 65 | 53 | 3 | 8 | 0 | 0 | 0 | 0 |
| -ve unfamiliar | 0 | 6 | 11 | 25 | 25 | 66 | 50 | 10 | 6 | 3 | 2 | 1 | 0 |
| +ve familiar | 1 | 8 | 7 | 22 | 29 | 56 | 62 | 15 | 1 | 0 | 0 | 0 | 0 |
| +ve same | 0 | 1 | 14 | 26 | 25 | 65 | 66 | 9 | 1 | 0 | 0 | 0 | 0 |
| +ve unfamiliar | 0 | 2 | 10 | 22 | 36 | 71 | 46 | 10 | 5 | 1 | 0 | 0 | 0 |

**Side Effects from Previous COVID Vaccines*Condition**

| | first dose | second dose | both doses | no side effects |
|---|---|---|---|---|
| -ve familiar | 61 | 28 | 21 | 93 |
| -ve same | 61 | 26 | 17 | 99 |
| -ve unfamiliar | 62 | 24 | 26 | 93 |
| +ve familiar | 57 | 20 | 22 | 102 |
| +ve same | 65 | 23 | 23 | 96 |
| +ve unfamiliar | 66 | 20 | 21 | 96 |

### Supplementary Materials 7

**Full statistical model for the primary outcome (including interactions between the factors (Framing/Familiarity) and the planned covariate (Baseline Booster Intention))**

Variables are:

- intent_fv_t2 (primary outcome: booster intention (intent) for the framed vaccine (fv) post-manipulation (t2))
- intent_fv_t1 (covariate: booster intention (intent) for the framed vaccine (fv) at baseline (t1))

**Full model accounting for interactions with the covariate**

| | Sum Sq | Df | F value | Pr(>F) |
|---|---|---|---|---|
| (Intercept) | 48462.8455 | 1 | 173.6635 | 0.0000 |
| intent_fv_t1 | 543030.3892 | 1 | 1945.9146 | 0.0000 |
| Framing | 25.7614 | 1 | 0.0923 | 0.7613 |
| Familiarity | 4466.3893 | 2 | 8.0025 | 0.0004 |
| intent_fv_t1:Framing | 76.6206 | 1 | 0.2746 | 0.6004 |
| intent_fv_t1:Familiarity | 5867.6007 | 2 | 10.5131 | 0.0000 |
| Framing:Familiarity | 6002.1555 | 2 | 10.7542 | 0.0000 |
| intent_fv_t1:Framing:Familiarity | 4268.1448 | 2 | 7.6473 | 0.0005 |
| Residuals | 337664.7592 | 1210 | NA | NA |

| | eta.sq | eta.sq.part |
|---|---|---|
| intent_fv_t1 | 0.5321 | 0.6166 |
| Framing | 0.0000 | 0.0001 |
| Familiarity | 0.0044 | 0.0131 |
| intent_fv_t1:Framing | 0.0001 | 0.0002 |
| intent_fv_t1:Familiarity | 0.0057 | 0.0171 |
| Framing:Familiarity | 0.0059 | 0.0175 |
| intent_fv_t1:Framing:Familiarity | 0.0042 | 0.0125 |

**Full model with pre-registered contrasts for Vaccine Familiarity**

- cont1 = Same Vaccine vs. Other Vaccines [Familiar and Unfamiliar combined]
- cont2 = Familiar Vaccine vs. Unfamiliar Vaccine

| | Sum Sq | Df | F value | Pr(>F) |
|---|---|---|---|---|
| (Intercept) | 48462.8455 | 1 | 173.6635 | 0.0000 |
| intent__fv__t1 | 543030.3892 | 1 | 1945.9146 | 0.0000 |
| Framing | 25.7614 | 1 | 0.0923 | 0.7613 |
| cont1 | 3134.4070 | 1 | 11.2319 | 0.0008 |
| cont2 | 1335.9564 | 1 | 4.7873 | 0.0289 |
| intent__fv__t1:Framing | 76.6206 | 1 | 0.2746 | 0.6004 |
| intent__fv__t1:cont1 | 4420.8055 | 1 | 15.8417 | 0.0001 |
| Framing:cont1 | 1413.4722 | 1 | 5.0651 | 0.0246 |
| intent__fv__t1:cont2 | 1553.5052 | 1 | 5.5669 | 0.0185 |
| Framing:cont2 | 4593.6316 | 1 | 16.4610 | 0.0001 |
| intent__fv__t1:Framing:cont1 | 1225.7388 | 1 | 4.3924 | 0.0363 |
| intent__fv__t1:Framing:cont2 | 3122.1111 | 1 | 11.1879 | 0.0008 |
| Residuals | 337664.7592 | 1210 | NA | NA |

| | eta.sq | eta.sq.part |
|---|---|---|
| intent__fv__t1 | 0.5321 | 0.6166 |
| Framing | 0.0000 | 0.0001 |
| cont1 | 0.0031 | 0.0092 |
| cont2 | 0.0013 | 0.0039 |
| intent__fv__t1:Framing | 0.0001 | 0.0002 |
| intent__fv__t1:cont1 | 0.0043 | 0.0129 |
| Framing:cont1 | 0.0014 | 0.0042 |
| intent__fv__t1:cont2 | 0.0015 | 0.0046 |
| Framing:cont2 | 0.0045 | 0.0134 |
| intent__fv__t1:Framing:cont1 | 0.0012 | 0.0036 |
| intent__fv__t1:Framing:cont2 | 0.0031 | 0.0092 |

### Supplementary Materials 8

##### Pre-registered ANCOVA (without interactions between factors and covariate)

The following presents the results from the pre-registered ANCOVA that did not account for interactions between the covariate (baseline booster intention) and the Framing/Familiarity factors.

##### Main Effects Model

| | Sum Sq | Df | F value | Pr(>F) |
|---|---|---|---|---|
| (Intercept) | 81587.3803 | 1 | 285.3356 | 0.0000 |
| intent_fv_t1 | 607341.4178 | 1 | 2124.0559 | 0.0000 |
| Framing | 667.2802 | 1 | 2.3337 | 0.1269 |
| Familiarity | 820.2857 | 2 | 1.4344 | 0.2387 |
| Framing:Familiarity | 1987.1180 | 2 | 3.4748 | 0.0313 |
| Residuals | 347410.7352 | 1215 | NA | NA |

| | eta.sq | eta.sq.part |
|---|---|---|
| intent_fv_t1 | 0.5951 | 0.6361 |
| Framing | 0.0007 | 0.0019 |
| Familiarity | 0.0008 | 0.0024 |
| Framing:Familiarity | 0.0019 | 0.0057 |

##### Model with pre-registered contrasts for Vaccine Familiarity

| | Sum Sq | Df | F value | Pr(>F) |
|---|---|---|---|---|
| (Intercept) | 81587.3803 | 1 | 285.3356 | 0.0000 |
| intent_fv_t1 | 607341.4178 | 1 | 2124.0559 | 0.0000 |
| Framing | 667.2802 | 1 | 2.3337 | 0.1269 |
| cont1 | 801.2642 | 1 | 2.8023 | 0.0944 |
| cont2 | 16.7860 | 1 | 0.0587 | 0.8086 |
| Framing:cont1 | 236.0587 | 1 | 0.8256 | 0.3637 |
| Framing:cont2 | 1747.3457 | 1 | 6.1110 | 0.0136 |
| Residuals | 347410.7352 | 1215 | NA | NA |

| | eta.sq | eta.sq.part |
|---|---|---|
| intent_fv_t1 | 0.5951 | 0.6361 |
| Framing | 0.0007 | 0.0019 |
| cont1 | 0.0008 | 0.0023 |
| cont2 | 0.0000 | 0.0000 |
| Framing:cont1 | 0.0002 | 0.0007 |
| Framing:cont2 | 0.0017 | 0.0050 |

**Framing*Familiarity Means**

| Framing | Familiarity | emmean | SE | df | lower.CL | upper.CL |
|---|---|---|---|---|---|---|
| Negative Frame | Unfamiliar | 77.0667 | 1.1868 | 1215 | 74.7383 | 79.3951 |
| Positive Frame | Unfamiliar | 82.0994 | 1.1939 | 1215 | 79.7572 | 84.4417 |
| Negative Frame | Familiar | 79.7128 | 1.1885 | 1215 | 77.3810 | 82.0445 |
| Positive Frame | Familiar | 78.8776 | 1.1952 | 1215 | 76.5327 | 81.2226 |
| Negative Frame | Same | 81.1027 | 1.1989 | 1215 | 78.7505 | 83.4549 |
| Positive Frame | Same | 81.3391 | 1.1946 | 1215 | 78.9955 | 83.6827 |

### Supplementary Materials 9

##### The effect of positive framing on vaccine switches

##### Full model including previous vaccine type ("prevax_type")

| | Sum Sq | Df | F value | Pr(>F) |
|---|---|---|---|---|
| (Intercept) | 48152.2239 | 1 | 173.6703 | 0.0000 |
| intent_fv_t1 | 466284.3106 | 1 | 1681.7448 | 0.0000 |
| Framing | 68.7237 | 1 | 0.2479 | 0.6187 |
| Familiarity | 4162.4448 | 2 | 7.5063 | 0.0006 |
| prevax_type | 1763.3901 | 1 | 6.3600 | 0.0118 |
| intent_fv_t1:Framing | 143.0262 | 1 | 0.5159 | 0.4728 |
| intent_fv_t1:Familiarity | 5531.5062 | 2 | 9.9752 | 0.0001 |
| Framing:Familiarity | 7062.0571 | 2 | 12.7353 | 0.0000 |
| intent_fv_t1:prevax_type | 945.5318 | 1 | 3.4102 | 0.0650 |
| Framing:prevax_type | 7.3229 | 1 | 0.0264 | 0.8709 |
| Familiarity:prevax_type | 207.0593 | 2 | 0.3734 | 0.6885 |
| intent_fv_t1:Framing:Familiarity | 5204.8395 | 2 | 9.3861 | 0.0001 |
| intent_fv_t1:Framing:prevax_type | 0.3171 | 1 | 0.0011 | 0.9730 |
| intent_fv_t1:Familiarity:prevax_type | 317.3683 | 2 | 0.5723 | 0.5644 |
| Framing:Familiarity:prevax_type | 1611.3401 | 2 | 2.9058 | 0.0551 |
| intent_fv_t1:Framing:Familiarity:prevax_type | 930.3705 | 2 | 1.6778 | 0.1872 |
| Residuals | 332160.1508 | 1198 | NA | NA |

| | eta.sq | eta.sq.part |
|---|---|---|
| intent_fv_t1 | 0.4569 | 0.5840 |
| Framing | 0.0001 | 0.0002 |
| Familiarity | 0.0041 | 0.0124 |
| prevax_type | 0.0017 | 0.0053 |
| intent_fv_t1:Framing | 0.0001 | 0.0004 |
| intent_fv_t1:Familiarity | 0.0054 | 0.0164 |
| Framing:Familiarity | 0.0069 | 0.0208 |
| intent_fv_t1:prevax_type | 0.0009 | 0.0028 |
| Framing:prevax_type | 0.0000 | 0.0000 |
| Familiarity:prevax_type | 0.0002 | 0.0006 |
| intent_fv_t1:Framing:Familiarity | 0.0051 | 0.0154 |
| intent_fv_t1:Framing:prevax_type | 0.0000 | 0.0000 |
| intent_fv_t1:Familiarity:prevax_type | 0.0003 | 0.0010 |
| Framing:Familiarity:prevax_type | 0.0016 | 0.0048 |
| intent_fv_t1:Framing:Familiarity:prevax_type | 0.0009 | 0.0028 |

#### Vaccine Switch: AstraZeneca to Pfizer and Moderna

| | Sum Sq | Df | F value | Pr(>F) |
|---|---|---|---|---|
| (Intercept) | 34492.1950 | 1 | 105.8593 | 0.0000 |
| intent_fv_t1 | 139372.9781 | 1 | 427.7470 | 0.0000 |
| Framing | 11.5190 | 1 | 0.0354 | 0.8510 |
| Familiarity | 833.9037 | 1 | 2.5593 | 0.1104 |
| intent_fv_t1:Framing | 7.8985 | 1 | 0.0242 | 0.8764 |
| intent_fv_t1:Familiarity | 1172.9565 | 1 | 3.5999 | 0.0585 |
| Framing:Familiarity | 4799.7488 | 1 | 14.7308 | 0.0001 |
| intent_fv_t1:Framing:Familiarity | 3707.3265 | 1 | 11.3781 | 0.0008 |
| Residuals | 130983.8324 | 402 | NA | NA |

| | eta.sq | eta.sq.part |
|---|---|---|
| intent_fv_t1 | 0.5019 | 0.5155 |
| Framing | 0.0000 | 0.0001 |
| Familiarity | 0.0030 | 0.0063 |
| intent_fv_t1:Framing | 0.0000 | 0.0001 |
| intent_fv_t1:Familiarity | 0.0042 | 0.0089 |
| Framing:Familiarity | 0.0173 | 0.0353 |
| intent_fv_t1:Framing:Familiarity | 0.0134 | 0.0275 |

#### Vaccine Switch: Pfizer to Moderna

| | Sum Sq | Df | F value | Pr(>F) |
|---|---|---|---|---|
| (Intercept) | 26187.1494 | 1 | 59.7228 | 0.0000 |
| intent_fv_t1 | 111498.8315 | 1 | 254.2860 | 0.0000 |
| Framing | 1744.2603 | 1 | 3.9780 | 0.0475 |
| intent_fv_t1:Framing | 791.0549 | 1 | 1.8041 | 0.1808 |
| Residuals | 86818.6664 | 198 | NA | NA |

| | eta.sq | eta.sq.part |
|---|---|---|
| intent_fv_t1 | 0.5565 | 0.5622 |
| Framing | 0.0087 | 0.0197 |
| intent_fv_t1:Framing | 0.0039 | 0.0090 |

**Model with pre-registered contrasts for Vaccine Familiarity**

| | Sum Sq | Df | F value | Pr(>F) |
|---|---|---|---|---|
| (Intercept) | 4.533390e+09 | 1 | 29.5304 | 0.0000 |
| Framing | 7.457785e+06 | 1 | 0.0486 | 0.8256 |
| cont1 | 2.443971e+08 | 1 | 1.5920 | 0.2073 |
| cont2 | 2.947225e+08 | 1 | 1.9198 | 0.1661 |
| Framing:cont1 | 5.626220e+04 | 1 | 0.0004 | 0.9847 |
| Framing:cont2 | 1.195929e+08 | 1 | 0.7790 | 0.3776 |
| Residuals | 1.866752e+11 | 1216 | NA | NA |

| | eta.sq | eta.sq.part |
|---|---|---|
| Framing | 0.0000 | 0.0000 |
| cont1 | 0.0013 | 0.0013 |
| cont2 | 0.0016 | 0.0016 |
| Framing:cont1 | 0.0000 | 0.0000 |
| Framing:cont2 | 0.0006 | 0.0006 |

**Mean estimates * Framing**

| Framing | emmean | SE | df | lower.CL | upper.CL |
|---|---|---|---|---|---|
| Negative Frame | 2004.294 | 501.2572 | 1216 | 1020.8692 | 2987.719 |
| Positive Frame | 1848.045 | 501.2893 | 1216 | 864.5571 | 2831.533 |

### Supplementary Materials 10

##### Post-hoc Familiarity * Framing * Baseline interactions among those with scores above and below the mean of baseline booster intention

###### Side Effect Severity

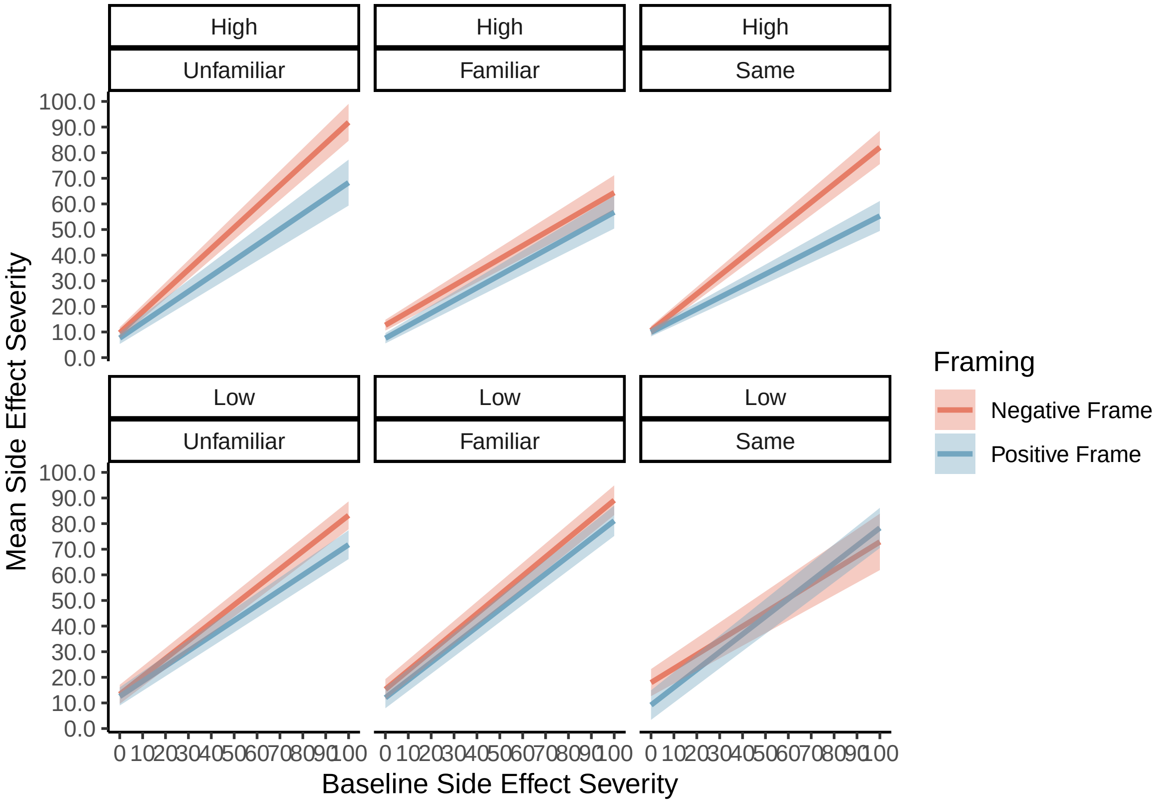

##### Booster Acceptance: Satisfaction and Happiness

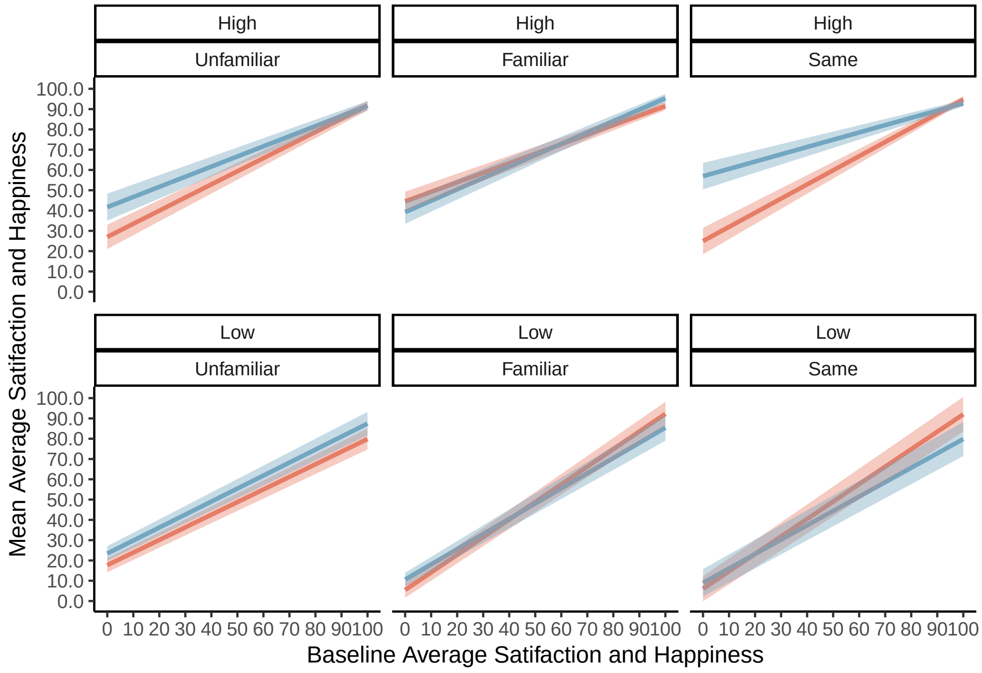


##### Booster Acceptance: Anxiety

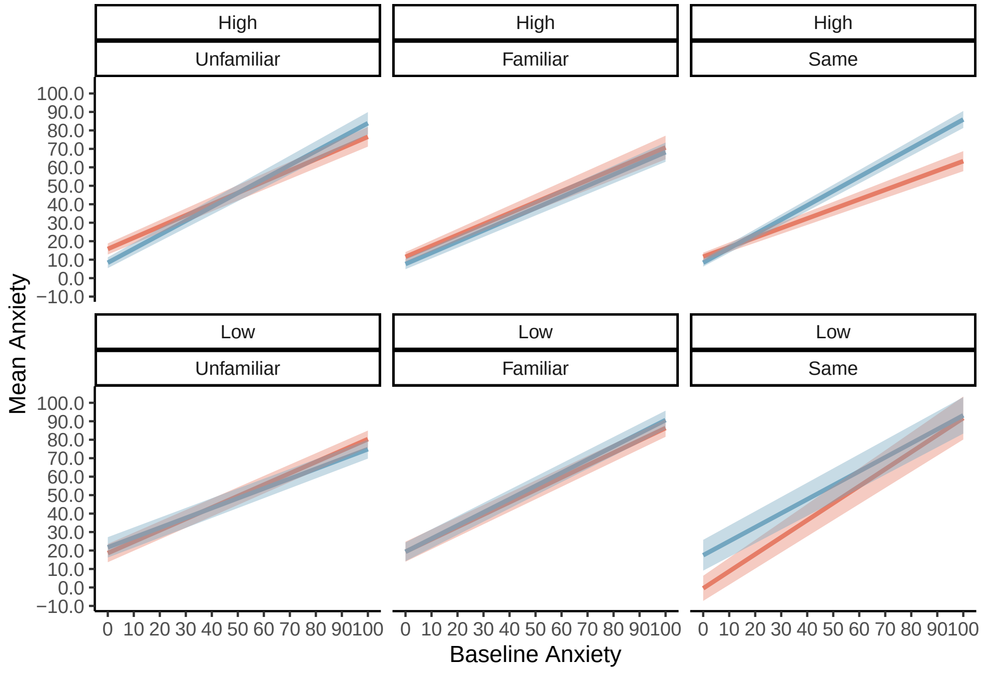
